# Targeted deep brain stimulation of the motor thalamus improves speech and swallowing motor functions after cerebral lesions

**DOI:** 10.1101/2024.09.16.24312391

**Authors:** Erinn M. Grigsby, Lilly W. Tang, Arianna Damiani, Jonathan C. Ho, Isabella M. Montanaro, Sirisha Nouduri, Sara Trant, Theodora Constantine, Gregory M. Adams, Kevin Franzese, Bradford Z. Mahon, Julie A. Fiez, Donald J. Crammond, Kaila L. Stipancic, Jorge A. Gonzalez-Martinez, Elvira Pirondini

## Abstract

Speech and swallowing are complex motor acts that depend upon the integrity of input neural signals from motor cortical areas to control muscles of the head and neck. Lesions damaging these neural pathways result in weakness of key muscles causing dysarthria and dysphagia, leading to profound social isolation and risk of aspiration and suffocation. Here we show that Deep Brain Stimulation (DBS) of the motor thalamus improved speech and swallowing functions in two participants with dysarthria and dysphagia. First, we proved that DBS increased excitation of the face motor cortex, augmenting motor evoked potentials, and range and speed of motion of orofacial articulators in n=10 volunteers with intact neural pathways. Then, we demonstrated that this potentiation led to immediate improvement in swallowing functions in a patient with moderate dysphagia and profound dysarthria as a consequence of a traumatic brain lesion. In this subject and in another with mild dysarthria, we showed that DBS immediately ameliorated impairments of respiratory, phonatory, resonatory, and articulatory control thus resulting in a clinically significant improvement in speech intelligibility. Our data provide first-in-human evidence that DBS can be used to treat dysphagia and dysarthria in people with cerebral lesions.

---

Natural and intelligible speech requires control of four subsystems: respiration, phonation, resonance, and articulation; similarly, swallowing involves sequential orchestration of movements from the oral cavity, pharynx, larynx, and esophagus to safely and efficiently ingest substances into the stomach. Precise and coordinated activation of these systems depends upon the integrity of both the cortico-spinal tract (CST), which innervates the muscles of respiration located in the thorax, neck, and shoulder, and the cortico-bulbar tract (CBT), which provides the bilateral innervation of laryngeal, palatal, lingual, and facial muscles^1^. Subcortical lesions that interrupt any of these tracts, as a consequence of stroke, traumatic brain injury (TBI), brain tumors, or neurodegenerative disorders, can cause weakness and deficits in the volitional control of facial and oropharyngeal muscles. This might result in a variety of undesirable auditory-perceptual speech characteristics, such as voice breaks and quality impairments, reduced speech intensity or imprecision of sound production. Any of these impairments alone, or in combination, can contribute to reduced speech quality and intelligibility, as a result of dysarthria^1–4^. Additionally, loss of oropharyngeal control can lead to dysphagia, i.e., swallowing difficulties, requiring alternative means of nutrition and increasing the risk of aspiration and pulmonary complications^5^. There are currently around 5 million people living with these conditions in the United States alone and these numbers are expected to escalate with the increase in life expectancy^6–8^.

Regardless of the etiology, speech therapy remains the standard treatment for dysarthria and dysphagia, where speech-language pathologists (SLPs) introduce compensatory behavioral techniques to improve speech and swallowing functions^9^. Yet the efficacy of these methods relies on individual effort and retention both during and after the course of the therapy^10–12^. External augmentative or substitutive communication devices^13^ can palliate unsuccessful behavioral interventions, but these methods do not restore natural speech production, leaving patients with difficulty communicating effectively, resulting in greater isolation and depression^14–17^. Additionally, without successful remediation of dysphagia, inability to safely feed persists, leading to accelerated deterioration of health^14–17^.

In most cases, damage to the CST and CBT is incomplete. Yet, the residual excitatory descending fibers are insufficient to fully activate the lower motoneurons, leading to functional deficits. Facilitating the activation of the residual axons could reestablish the missing excitation, restoring speech and swallowing motor functions^18^. This facilitation could be achieved by increasing the excitability of cortical neurons particularly in the face representation of the motor cortex, thereby increasing CST and CBT motor output and, consequently, enhancing the facial and oropharyngeal movements responsible for speech production and swallowing. In this regard, a few previous works proposed the use of non-invasive neuromodulation interventions, such as transcranial magnetic stimulation (TMS) and transcranial direct current stimulation (tDCS), to increase cortical excitability and improve dysarthria and dysphagia, yet with mixed results^19–23^.

We recently demonstrated that electrical stimulation of direct excitatory connections to the motor cortex can increase the excitability of cortico-spinal motoneuron, potentiating upper-limb motor output^18^. Specifically, with intra-operative experiments in non-human primates and human subjects, we showed that low frequency (50-100Hz) deep brain stimulation (DBS) of the motor thalamus enhanced the excitability of the CST by recruiting afferent fibers to the motor cortex. This increased excitability improved arm and hand strength and volitional control in a participant with a severe paralysis as a consequence of TBI. Importantly, these effects appeared immediately when switching ON the stimulation and vanished when the stimulation was turned OFF. These results were paralleled by a recent study showing that, in stroke patients, long-term increased cortical excitability by DBS of the cerebellar dentate nucleus resulted in upper-limb motor improvements that outperformed results obtained with TMS^24^. These promising effects could be due to the higher selectivity and continuous nature of DBS^25–27^. Paralleling these findings and in relation to speech and swallowing functions, a few recent studies have shown that low frequency DBS of the subthalamic nuclei could improve speech intelligibility and aspiration in Parkinson’s disease (PD) patients suffering degenerative hypophonia and dysarthria. Yet, no previous studies attempted to use DBS to treat dysarthria and dysphagia in subjects with damaged descending motor fibers as a consequence of cerebral lesions.

Importantly, the motor thalamus also has direct excitatory connections to the agranular motor cortical areas representing the face and larynx^28–38^. We therefore hypothesized that electrical stimulation of the motor thalamus could immediately increase excitability of the CBT improving control of facial and oropharyngeal muscles. Our preliminary results in non-human primates supported our hypothesis^18^. Indeed, stimulation of the motor thalamus induced potentiation of motor evoked potentials (MEPs) of orofacial muscles.

Building on this previous evidence, here we reasoned that targeted DBS of the motor thalamus could be tailored to improve the activation and control of facial and oropharyngeal muscles, immediately lessening swallowing and speech motor deficits. For this, we first demonstrated that electrical stimulation of the motor thalamus potentiated MEPs of facial and oropharyngeal muscles in human subjects (n=7). This potentiation resulted in an increase in range of motion and speed of facial articulators during movements requiring control of lips, jaw, and tongue in n = 3 human subjects with intact descending motor fibers. Importantly, these effects were also present in a subject with a severe chronic traumatic lesion of the CST and CBT, which led to profound dysarthria and moderate dysphagia. Additionally, stimulation improved swallowing functions evaluated with a videofluoroscopic swallow study. We then demonstrated, in the same participant and in a second subject suffering from mild dysarthria due to TBI, that thalamic stimulation ameliorated control of the four speech subsystems (respiration, phonation, resonance, and articulation), leading to an immediate and clinically significant improvement in speech intelligibility and quality as assessed by blinded SLPs and naive listeners.

## Results

### Stimulation of the motor thalamus potentiates motor evoked potentials of facial and oropharyngeal muscles

We previously demonstrated that stimulation of the motor thalamus increases primary motor cortex (M1) excitability and upper-limb motor output via the CST. Specifically, we showed that when electrically stimulating the motor thalamus, the amplitude of M1 direct cortical stimulation (DCS)-MEPs of contralateral upper-limb muscles was immediately increased both in humans and monkeys^18^ paralleling previous studies^28,39^. Interestingly, in monkeys, we also found a significant potentiation of contralateral facial and oropharyngeal muscle MEPs. Here we first sought to demonstrate that targeting the motor thalamus will cause a similar potentiation in facial and oropharyngeal muscles in human subjects (**Fig. 1a**). To test this, we leveraged intraoperative electrophysiological recordings and stimulation tests routinely performed in patients undergoing DBS implantation (n=7, subjects S01-S07, 3 males, 4 females, **Supplementary Table 1**). These experiments followed a protocol consistent with our previous study on the mechanism of motor thalamus stimulation^18^. Specifically, we placed bilateral sub-cutaneous EMG needle electrodes in up to four facial and oropharyngeal muscles typically associated with speech and swallowing physiology (masseter, orbicularis oris, mylohyoid, and cricothyroid, **Fig. 1a-b**). We then implanted a 6-channel subdural strip electrode (Adtech, Oak Creek, WI, USA) over the face representation of the primary motor cortex (M1). To verify electrode position, we performed DCS of the subdural electrode and recorded facial DCS-MEPs. We selected the optimal subdural stimulation contact as the contact that elicited the largest MEPs and recruited the greatest number of contralateral facial and oropharyngeal muscles.

**Fig. 1.**
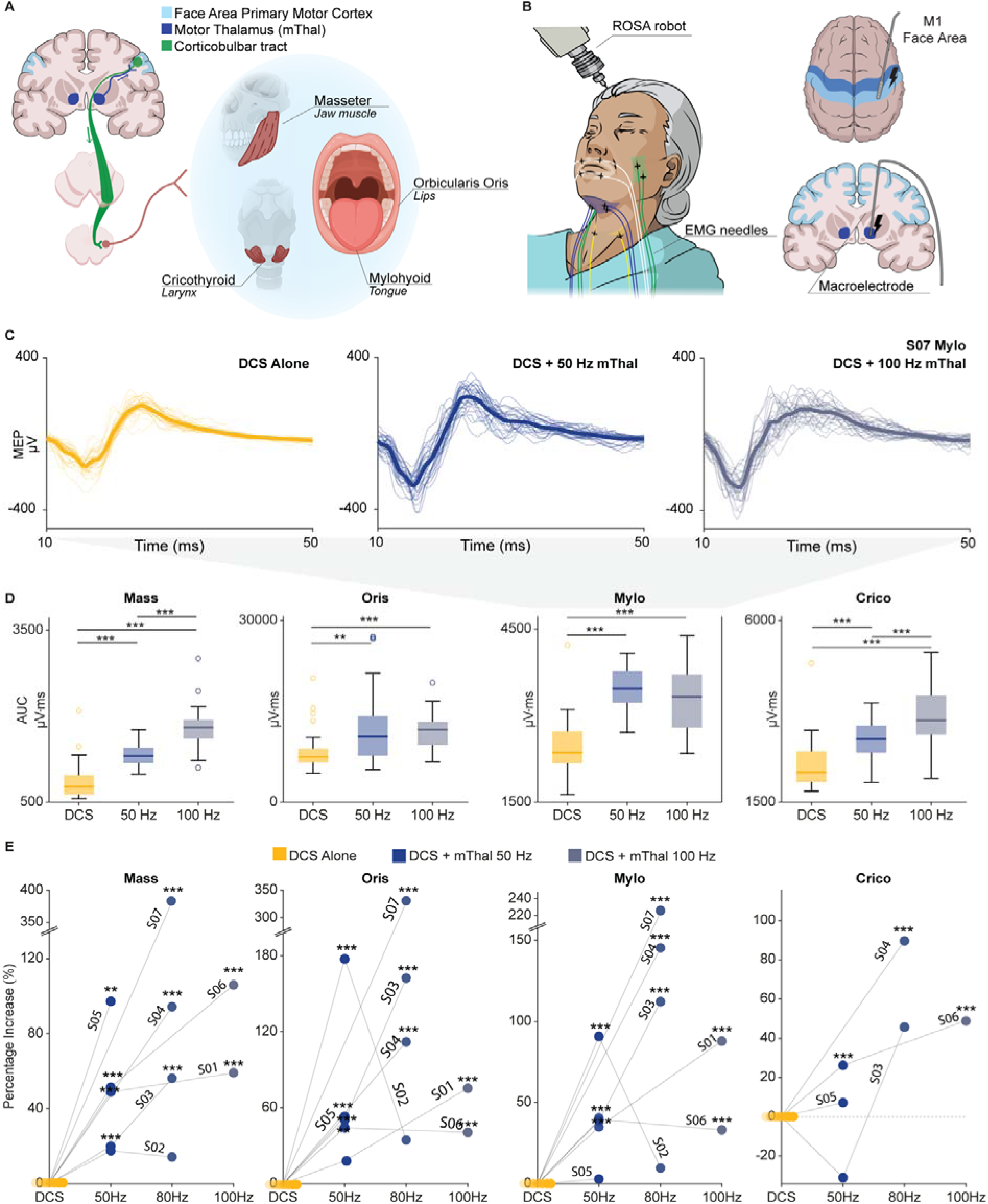
Motor thalamus stimulation amplifies facial and oropharyngeal motor evoked potentials in humans. **(a)** Schematic of the mechanisms of action. The motor thalamus (mThal; dark blue) projects excitatory connections to the face representation of the primary motor cortex (M1, light blue). Electrical stimulation of the motor thalamus increases excitation of the face motor cortex consequently potentiating muscles associated with the main facial articulators involved in swallowing and speech, jaw (masseter), lips (orbicularis oris), tongue (mylohyoid), and larynx (cricothyroid), via projections through the CBT (green) and the CST (not represented). **(b)** Experimental setup for the intraoperative experiments. Needle electrodes were inserted into the masseter (Mass, green), the orbicularis oris (Oris, white), the mylohyoid (Mylo, blue), and the cricothyroid (Crico, yellow) muscles. A subdural strip electrode was placed over the facial area of M1. Microelectrodes traversed the motor thalamus. **(c)** Example MEP traces with M1-DCS alone and M1-DCS paired with motor thalamus stimulation at 50 and 100 Hz for subject S07’s mylohyoid muscle MEPs (n=30 traces for each condition). **(d)** Example box pots of area-under-the-curve (AUC) for the MEPs with and without stimulation of the motor thalamus for Mas, Oris, Mylo, and Crico muscles (n=30 traces for each condition and muscle) from subject S06. For all boxplots, the whiskers extend to the maximum spread excluding outliers. Central, top, and bottom lines represent the median, 25th, and 75th percentile, respectively. **(e)** Scatter plots of the percentage of increase of AUC of each facial and oropharyngeal muscle between DCS alone and DCS paired with motor thalamus stimulation at 50, 80, and 100 Hz for all n = 7 patients. For all panels, statistical significance was assessed with two-tailed bootstrapping with Bonferroni correction: p<0.05 (*), p<0.01 (**), p<0.001 (***).

We then recorded facial and oropharyngeal MEPs with M1-DCS alone or M1-DCS paired with motor thalamus stimulation at 50, 80, and 100 Hz from a macroelectrode (Neuro Omega, Alpha Omega Engineering, Israel) implanted in the same hemisphere (see **Supplementary Table 2** for individual parameters of stimulation). Consistent with our previous results for upper-limb and orofacial muscles in monkeys^18^, we observed an immediate significant increase in the MEP amplitudes of contralateral facial and oropharyngeal muscles when M1-DCS was paired with motor thalamus stimulation compared to DCS alone (i.e., 28-330% increase over all recorded muscles and tested frequencies, **Fig. 1c-d** and **Extended Data Fig. 1a**). Interestingly, the increase was present for all recorded muscles, which were associated with the main facial articulators involved in swallowing and speech: jaw (masseter), lips (orbicularis oris), tongue (mylohyoid), and larynx (cricothyroid).

Because speech requires bilateral muscle control, and CBT from one hemisphere innervates muscles on both sides of the face, we also explored the effects of motor thalamus stimulation on the ipsilateral facial and oropharyngeal muscles (i.e., muscles on the same side of DCS and motor thalamus stimulation)^1^. As hypothesized, there was an immediate and significant potentiation of the ipsilateral facial and oropharyngeal MEPs when M1-DCS was paired with stimulation of the motor thalamus (**Extended Data Fig. 1a**), though potentiation was significantly stronger for contralateral muscles (i.e., on average over all muscles and participants contralateral potentiation was 83% greater than it was for ipsilateral counterparts, **Extended Data Figure 1b**).

In summary, our results demonstrate that targeted electrical stimulation of motor thalamus increases excitability of CST and CBT enhancing DCS-MEPs of facial and oropharyngeal muscles in human subjects with intact descending fibers. These results were consistent with our previous findings of facilitation of upper-limb DCS-MEPs, further supporting our proposed mechanisms of action of motor thalamus DBS.

### Stimulation of the motor thalamus increases speed and range of motion of facial articulators during face movements

We then sought to demonstrate whether the observed potentiation of DCS-MEP amplitudes would result in augmentation of motor output during overt voluntary facial movements and with motor thalamus stimulation. Because of the inherent challenges associated with behavioral testing intraoperatively, to test for this, we performed stimulation experiments in three human subjects (2 male, 1 female) who underwent surgical implantation of stereo-electroencephalography (sEEG) electrodes for seizure localization. The sEEG electrode placement for these patients included extended coverage of the motor thalamus. Tests were performed during patient hospitalization in the epilepsy monitoring unit. The subjects were instructed to complete facial motor tasks, rapidly moving between instructed (tongue in/out, smile, mouth open/close, and lip pucker) and neutral facial expressions at a comfortable pace, with and without motor thalamus stimulation (see **Supplementary Table 3** for individual parameters of stimulation). Importantly, these experiments allowed us to stimulate the motor thalamus while the patients had full autonomy over their voluntary facial motor control. Indeed, all three participants had intact CBT and no clinical history of swallowing, speech, or facial motor deficits. Surface-electrode EMGs from the masseter, orbicularis oris, and mylohyoid were collected in two subjects (S08 and S10) to assess activation of orofacial muscle (**Fig. 2a**).

**Fig. 2.**
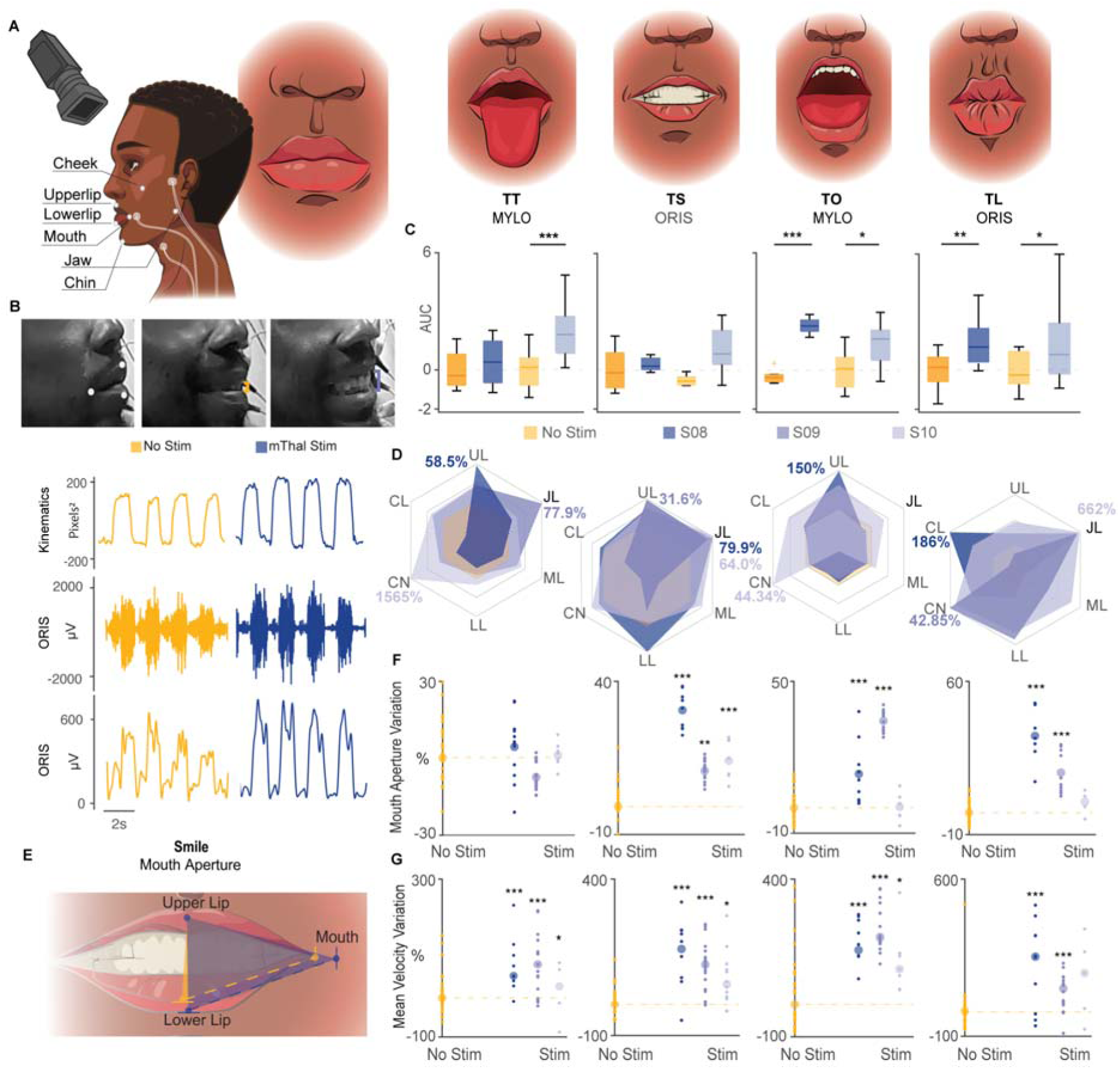
Motor thalamus stimulation enhances voluntary facial movements in humans with intact descending fibers. **(a)** *Left:* Behavioral experiment set up for the tests during facial movements with highlighted surface EMG electrode placement and facial articulators. *Right:* Voluntary facial tasks alternating between neutral facial expression (left panel) and tongue in/out (TT), smile (TS), mouth open/close (TO), or lip pucker (TL) facial expressions. **(b)** *Top:* Frame captured from videos showing enhanced facial movement of TS in subject S08. *Bottom:* Kinematic trace of cumulative movement amplitude between mouth, upper lip and lower lip articulators. Raw and normalized EMG trace of orbicularis oris. *Left*, without stimulation (yellow); *right*, with motor thalamus stimulation (blue). **(c)** Boxplots of z-score of EMG AUC for mylohyoid and orbicularis oris during facial tasks (n=2) without (yellow) and with (blue) stimulation. **(d)** Radial plots of normalized change in amplitude of kinematic facial articulators between no stimulation (yellow) and stimulation (blue). Different blue intensities correspond to different subjects (n=3). Articulators include upper lip (UL), jaw (JL), cheek (CL), mouth (ML), chin (CN), and lower lip (LL). **(e)** Example of mouth aperture calculated from 3 representative facial articulators for TS. **(f)** Percentage of variation in mouth aperture between no stimulation (yellow) and stimulation (blue) for n=3 subjects. Each point corresponds to one movement. **(g)** Percentage of variation in mean velocity of a representative facial articulator capturing the main direction of movement specific to each task, comparing stimulation OFF (yellow) and stimulation ON (blue) for n=3 subjects. Each point corresponds to one movement. For all boxplots, the whiskers extend to the maximum spread excluding outliers. Central, top, and bottom lines represent the median, 25th, and 75th percentile, respectively. For all panels, statistical significance was assessed with two-tailed bootstrapping with Bonferroni correction: p<0.05 (*), p<0.01 (**), p<0.001 (***).

As expected, we observed an immediate and significant enhancement in muscle activation with motor thalamus stimulation compared to the no thalamic stimulation condition (**Fig. 2b-c**). Importantly, the area-under-the-curve (AUC) of the voluntary EMG envelopes was significantly larger with thalamic stimulation only when the participants were actively performing facial movements. Indeed, EMG activation at rest was similar between stimulation ON and OFF conditions (**Extended Data Figure 2**), suggesting that the observed potentiation was not induced by a direct activation of CST and CBT elicited by current spread from thalamic stimulation. In this regard, we previously demonstrated extensively the absence of current spreading to CST and CBT from thalamic stimulation in monkeys and human subjects (sees **Figure 4** and **7** and **Supplementary Figure 4** in Ho & Grigsby et al., 2024^18^). Additionally, no participants (S08-S10) experienced any discomfort or impairment of speech abilities during thalamic stimulation, reinforcing the safety and lack of adverse side-effects of our approach.

Videos of subjects completing the facial tasks were recorded for kinematic analysis. We assessed the amplitude of the movements of the main facial articulators involved in the motor tasks: jaw, cheek, mouth, upper and lower lip, and chin. As hypothesized, motor thalamus stimulation immediately and significantly increased movement amplitude as compared to no stimulation trials in all participants, paralleling the increased EMG activation (i.e., increase range between 31.6% and 1565% across all facial articulators and participants, **Fig. 2d, Video 1**). To further quantify this increase, the most representative three facial articulators for each task were then selected to calculate the mouth aperture, which was immediately and significantly increased with thalamic stimulation across participants for smile, mouth open/close, and lip pucker, but not for tongue in/out (**Fig. 2e** and **2f**). However, movement speed was significantly increased when the stimulation was turned ON compared to no stimulation for all four tasks (**Fig. 2g, Video 1**).

Overall, these results demonstrate that motor thalamus stimulation enhances voluntary facial motor output in humans with intact CST and CBT.

### Stimulation of the motor thalamus potentiates facial movements in a person with severe CBT lesion (TBI01)

Given the observed effects of motor thalamus stimulation on voluntary facial movement in humans with intact CBT, we next sought to assess whether stimulation of the motor thalamus could also potentiate facial movements in humans with partially lesioned CST and CBT. We had the opportunity to test the impact of motor thalamus stimulation in a patient who sustained bi-hemispheric partial traumatic subcortical injuries (TBI01), resulting in bilateral hemiparesis, as well as severe facial paresis, moderate dysphagia, and profound dysarthria. The patient was implanted with a chronic DBS system in the motor thalamus for clinical reasons. Diffuse tractography of his descending motor fibers confirmed the presence of bilateral lesions of the CBT, with more damage localized to the left side paralleling his exhibited motor symptoms (**Extended Data Figure 3a**).

We tested TBI01 over three sessions in which he was instructed to perform the same voluntary facial motor tasks completed by subjects S08-S10, with and without bilateral motor thalamus stimulation. For this study, thalamic stimulation was set to a frequency of 55 Hz only during the testing (see **Supplementary Table 3** for details on the stimulation parameters).

We recorded videos of TBI01 completing the tasks to perform quantitative kinematic analysis. Similar to subjects S08-S10, kinematic analysis showed an immediate and significant increase in amplitude and speed of facial movement when the stimulation was turned ON relative to no stimulation (**Fig. 3a-b** and **Video 2**). Indeed, we found a significant potentiation of movement amplitude (**Figure 3c**), mouth aperture (**Figure 3d-e**), and movement mean velocity (**Figure 3f**) during thalamic stimulation consistent over all days of testing (n = 3).

**Fig. 3.**
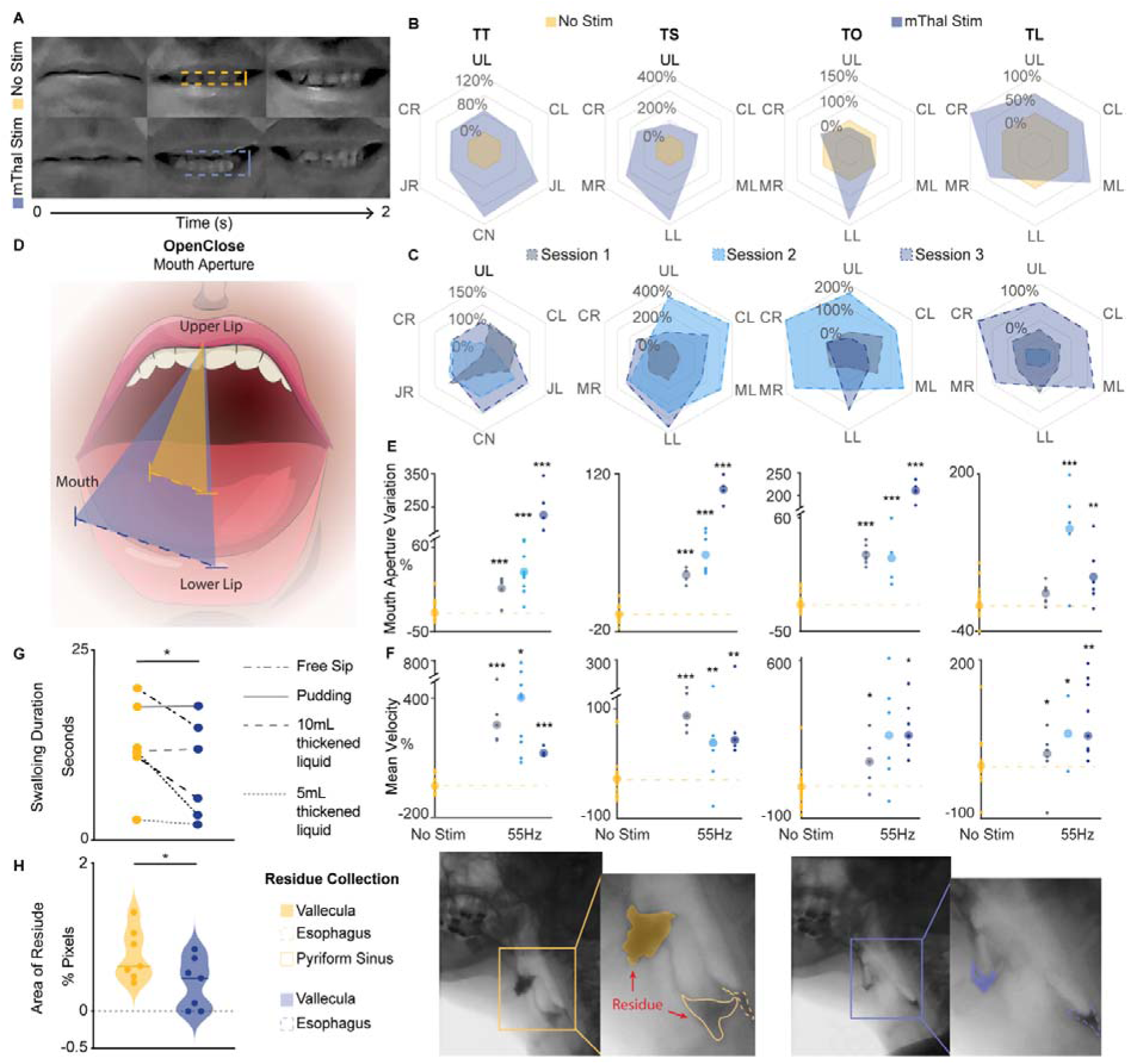
DBS improves facial movement and swallowing functions in a participant with a severe traumatic CBT lesion. **(a)** Frame captures from video showing TBI01 maximum range of motion of the lips during smile task for DBS OFF (yellow) and DBS ON (blue) conditions. **(b)** Radial plots of normalized change in amplitude of facial articulators during facial tasks between DBS OFF (yellow) and ON (blue) for session 1. Articulators include upper lip (UL), jaw left (JL), jaw right (JR), cheek left (CL), cheek right (CR), mouth left (ML), mouth right (MR), chin (CN), and lower lip (LL). **(c)** Radial plots of normalized change in amplitude of kinematic facial markers across three sessions of testing. Different blue intensities represent different sessions. **(d)** Mouth aperture calculated from 3 representative facial articulators for TO for session 1. **(e)** Percentage of variation in mouth aperture between DBS OFF (yellow) and DBS ON (blue) across sessions (n=3). Each point corresponds to one movement. **(f)** Percentage of variation in mean velocity of a representative facial articulator capturing the main direction of movement specific to each task between DBS OFF (yellow) and DBS ON (blue) across sessions (n=3). Each point corresponds to one movement. For panels e and f, statistical significance was assessed through two-tailed bootstrapping with Bonferroni correction: (*) p<0.05; (**) p<0.005; (***) p<0.001. **(g)** Total duration of various trials of the videofluoroscopic swallow study matched between DBS OFF and DBS ON. Please note that for 5 and 10mL thickened liquid trials, 2 repetitions were performed. **(h)** *Left:* Area of vallecula, esophagus, and pyriform sinus residues quantified from frame captures of phase-matched radiology video report across all trials. To be noted that with DBS ON there was no piriform sinus residue. For panels g and h, statistical significance was determined through one-tail parametric paired t-test, (*) p<0.05. *Right:* Frame captures of phase-matched radiology recordings of TBI01 videofluoroscopic swallow study for a representative trial of free sip of mildly thick liquid for DBS OFF (yellow) and DBS ON (blue). Residue bolus are shaded or outlined for reference.

Since participants perceive mild and transient paresthesia when the stimulation is turned ON, they cannot be blinded to the presence or absence of stimulation. Therefore, stimulation effects could be confounded by increased motivation of participant when they perceive stimulation is active. To account for this, we compared effects of stimulation at standard DBS frequency (130 Hz^25^) and low frequency (55 Hz) in TBI01, who was the only participant we could assess due to technical and time constraints (**Extended Data Figure 4a**). Importantly, TBI01 was unable to distinguish between the different frequencies. Stimulation at 130 Hz resulted in a general amplification of movement as compared to no stimulation. However, stimulation at 55 Hz showed greater amplification suggesting a stronger efficacy of the stimulation at lower frequencies (range of movement increased by 97% to 364.3% at 55 Hz versus a range of increase by 21% to 88.5% at 130 Hz, **Extended Data Figure 4b**).

Overall, these results demonstrate that motor thalamus potentiation of volitional control of facial movements is present in intact subjects and also in a patient with an injury of the descending motor fibers, indicating the possible clinical relevance of our approach.

### DBS improves swallowing functions in a participant with moderate dysphagia (TBI01)

We then investigated whether the effects of thalamic stimulation on potentiating facial movements would translate to a functional improvement in swallowing. Importantly, dysphagic patients show a delay in the transit of a liquid or solid bolus and/or an increase of residue during the oral, pharyngeal, or esophageal phase of swallowing due to loss of strength and coordination of facial and oropharyngeal muscles^40^. To assess these deficits and the effects of stimulation in TBI01, we conducted a videofluoroscopic swallow study^41^ (i.e., modified barium swallow test) with and without stimulation. A speech-language pathologist (SLP) who was blinded to the experimental design and stimulation conditions performed the clinical evaluation and interpretation of the study. Without stimulation, TBI01 presented with moderate oral dysphagia, mild pharyngeal dysphagia, and no deficits in the esophageal phase.

The SLP consistently reported improved oral and pharyngeal phase of swallowing with thalamic stimulation ON relative to the no stimulation condition (**Video 3**). She also reported reduced difficulty initiating volitional swallows and decreased oral residue when the stimulation was ON. However, with stimulation, there was an increased distention of the esophagus during the esophageal phase of swallowing. Importantly, this distention did not negatively affect the safety and efficiency of swallowing.

To further assess the effects of the stimulation, we quantified the duration of swallowing and the maximum amount of residue in the videofluoroscopic swallowing study. Paralleling the SLP report, the total duration of swallowing across all phases was significantly reduced when stimulation was ON as compared to the OFF condition (p = 0.038, n = 6 tasks), indicating a faster, more efficient swallow (**Fig. 3g**). Additionally, TBI01 had significantly less residue in the oral and pharyngeal phase of swallowing when stimulation was ON compared to no stimulation (p=0.037, n = 6 tasks), highlighting a positive effect of the stimulation particularly for deglutition of liquids (**Fig. 3h**).

These results collectively demonstrate the restorative effect of thalamic stimulation on enhancing facial and oropharyngeal muscle activity to improve swallowing function in patients with chronic lesions of the CBT.

### Stimulation of the motor thalamus improves respiratory, phonatory, resonatory, and articulatory control during speech in two participants with TBI

Inspired by the augmented facial and oropharyngeal muscle activation during voluntary motor tasks with thalamic stimulation, we sought to evaluate whether this increase would translate to an improvement in speech production in two participants with chronic speech deficits due to a traumatic CBT lesion (**Extended Data Fig. 3**). In addition to TBI01, who had profound dysarthria, we performed tests in a subject (TBI02) with mild dysarthria due to TBI who was implanted with temporary sEEG electrodes in the motor thalamus for seizure localization (**Supplementary Table 1**). Prior to any of the testing sessions, an SLP, who was blinded to the experimental design, performed a speech evaluation with both participants. TBI01 demonstrated a profound dysarthria characterized by impaired ability to control voice pitch or loudness and breath support for phonation, and imprecise articulation and distortions of consonants and vowels (most notably bilabial consonant sounds). TBI02 exhibited characteristics of mild dysarthria with limited distortion of consonant phonemes and increased imprecision with speech rate. During multiple sessions (**Supplementary Table 4** and **Extended Data Fig. 5**), we collected video and audio recordings as both participants completed a series of speech tests. Due to the significant difference in their deficits, we personalized the tests based on the participants’ speech ability. Specifically, we tasked TBI01 to repeat aloud single words five times. To assess the effects of the stimulation on different phonemes, we selected mono-to-triple syllable words that included the full spectrum of American English phonemes and required different levels of coordination and placement of lips, soft palette, and tongue (**Extended Data Fig. 5**). Instead, for TBI02 who suffered from milder deficits, we instructed him to repeat 2-word tongue-twisters as fast as possible for 20 seconds^42^. We selected the tongue-twisters to include word pairs that matched the words tested in TBI01 (**Extended Data Fig. 5**).

Since the speech deficits of TBI01 affected all four speech subsystems, we assessed effects of the stimulation separately on each of them. Vocal intensity was increased when the stimulation was turned ON as compared to stimulation OFF in 72.2% of the words (65.4% of which were statistically significant, **Fig. 4a**). Importantly, in 35.3% of these significant cases, the increase was above 5dB, which is a clearly noticeable change in loudness considered clinically relevant^43^, proving that thalamic stimulation was effective in improving breath support for volume control. Similarly, thalamic stimulation immediately and significantly reduced the number of voice breaks (mode: 1 break and 0 breaks, with stimulation OFF and ON, respectively, **Fig. 4b**) demonstrating an improvement in phonatory control. We further assessed effects of the stimulation on phonatory integrity by computing Cepstral Peak Prominence-smoothed (CPPS)^44,45^: a lower CPPS value is associated with a more dysphonic sound, often presenting as a hoarse, harsh, and/or strained voice quality. We observed a significant increase in the CPPS values when the stimulation was turned ON compared to no stimulation (median increase: 10.9%, **Fig. 4c**), further demonstrating a significant improvement in phonatory control. We also found an improvement in articulatory control. Indeed, participants suffering from dysarthria often experience deficits in control of the articulators, leading to an omission of consonants and distortion of vowels^1^. For this reason, we assessed articulatory control separately for consonants, by perceptually assessing presence/absence of each phoneme, and vowels, by quantifying changes in the main formants (i.e., changes in frequencies of F1 and F2 formants). Interestingly, we found that with stimulation ON there was an increase (24.4 ± 9.5%, **Fig. 4e**) in the perceptual presence of several consonant sounds (/b/, /m/, /f/, /d/, /l/, /g/, and /h/). Additionally, we found a change in the frequencies of the F1 formant, which encodes mouth control^46^ (increase for low vowels - /ei/, /Ɛ/ and /ɑ/ - and decrease for high vowels - /i/, /ɑɪ/, /ɝ/ and /oʊ/), and in the frequencies of F2 formant, which encodes tongue control^46^ (decrease for back vowels - /ɑ/, /ɔ/, and /ʌ/ - and an increase for front vowels - /eɪ/, /ɛ/, and /ɝ/ - **Fig. 4e**). These changes suggest an effect of the stimulation on mouth and tongue control consistent with the improvements found in the facial expression and swallowing tasks (**Fig. 2** and **3**). Finally, we assessed whether thalamic stimulation would reduce the amount of task duration-related fatigue that TBI01 would experience. For this, we computed the time required for TBI01 to complete repetition of the first five unique words and the last five unique words and compared it between stimulation ON and OFF. We reasoned that a participant would generally experience more fatigue at the end of a task than at the beginning of a task and would, therefore, required more time to complete productions at the end of the task as compared to the beginning of the task. Therefore, if stimulation decreased the task duration-related fatigue level, we would expect to observe larger differences in timing between the two conditions for the last five words. Our hypothesis was confirmed, i.e., the time to produce five words was statistically (p = 0.02) shorter for stimulation ON condition as compared to stimulation OFF only for the last five words, demonstrating that motor thalamus stimulation decreased fatigue level in TBI01 (**Fig. 4d**).

**Fig. 4.**
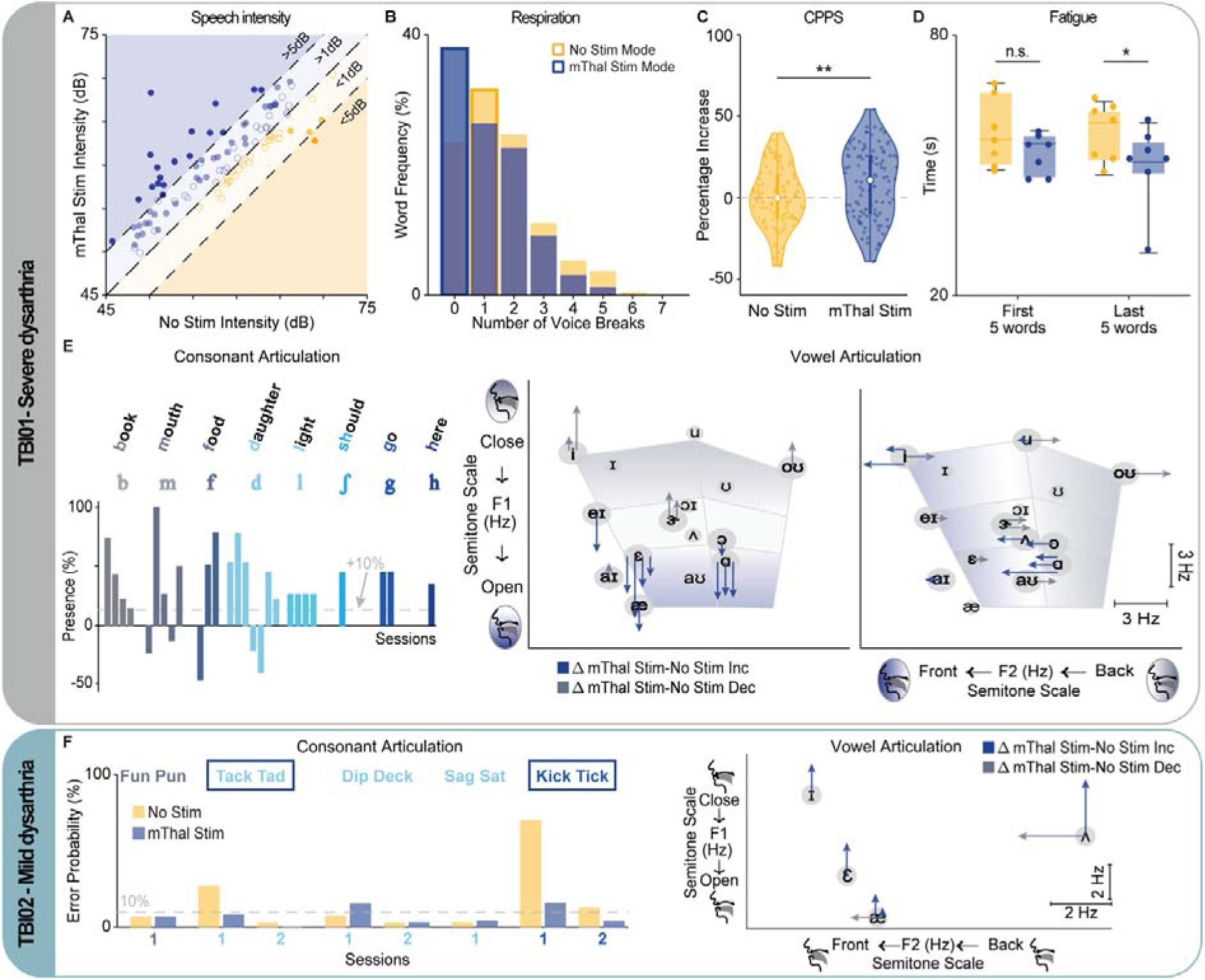
DBS improves respiratory, phonatory, resonatory, and articulatory control during speech in subjects with dysarthria. Results are separated by participants to capture improvements in severe dysarthria (TBI01, gray rectangle, panels a-e) and mild dysarthria (TBI02, teal rectangle, panel f). **(a)** Scatter plot of the speech intensity for the ON and OFF stimulation conditions for single words. Each dot (n = 108) represents a single word from a single session. In total we tested 52 unique words (see **Extended Data Figure 5**). Filled dots are statistically significant between stimulation ON and OFF (p<0.05). Dots and shades are colored based on the difference of mean intensity for the ON and OFF stimulation conditions (dark blue: Δ>5dB, blue: 0<=Δ<5dB, light yellow: −5dB<Δ<0, and gold: Δ<-5dB). **(b)** Histogram of the frequency of voice breaks for all words. The bars containing the mode values for stimulation OFF (mode = 1) and ON (mode = 0) are bolded. **(c)** Percentage increase in CPPS between stimulation OFF and ON across all sessions and words. The percent increase was calculated by normalizing all CPPS values by the median CPPS of all words during stimulation OFF conditions. **(d)** Boxplot of time duration necessary to complete repetitions of the first and last five words within a condition (~25 repetitions of single words) with stimulation OFF and ON. Statistical significance was assessed through parametric paired t-test across each session to standardize the number of words repeated within each condition (*) p<0.05. **(e)** *Left:* Percentage of perceptual presence for those consonants that reported a significant increase (>10%) in perceptual presence when the stimulation was ON as compared to stimulation OFF. Each bar represents a different session. All bars are normalized to their stimulation OFF average. The colors for the different phoneme match color in **Extended Data Figure 5**. *Right:* Arrows represent changes in formant frequencies during stimulation. Each arrow represents a session with a significant difference, with the length proportional to the change in frequency. Shown are all the vowels tested. **(f)** *Left:* Percentage decrease of errors during the “tongue twister” task with stimulation ON for TBI02. Blue box indicates tongue-twisters with error reductions > 10%. Tongue twisters are colored according to the first word’s consonant matching the colors in **Extended Data Figure 5**. *Right:* Arrows represent changes in formant frequencies during stimulation. Each arrow represents a session with a significant difference, with the length proportional to the change in frequency. Shown are all the vowels tested. For panels a, c, e, and f, statistical significance was assessed with two-tailed bootstrapping: p<0.05 (*), p<0.01 (**), p<0.001(***).

Similar to the facial expression tasks, to demonstrate that the effects of the stimulation were not confounded by motivation, we also assessed effects of stimulation at 130 Hz during the speech tasks for one day of recordings. While stimulation at 130 Hz and 55 Hz led to comparable values of speech intensity and CPPS (**Extended Data Figure 4c-d**), stimulation at 55 Hz produced better articulation. Indeed, overall stimulation at 130 Hz resulted in an increased perceptual presence of consonants as compared to no stimulation. Yet, for the majority of the consonants (7 out of 8) the increase was smaller than with stimulation at 55 Hz (11.7 ± 26.1% at 130 Hz whereas 45.7 ± 25.5% at 55Hz within the same session, **Extended Data Figure 4e**). Similarly, stimulation at 130 Hz led to smaller magnitude of changes and fewer significant changes in F1/F2 formats as compared to stimulation at 55 Hz (**Extended Data Figure 4f**).

Because TBI02 primarily demonstrated distortion of consonant phonemes and imprecision of phonation, we focused on evaluating the effect of stimulation on his articulatory control. For this, we first quantified the production errors in the tongue-twisters task, including *speech arrest* (e.g., struggling to initiate or complete words), *distorted or incorrect phoneme* (e.g., /tæk/ instead of /tɪk/), *slurring* words (e.g., prolonged consonants), and *stuttering* words (e.g., repetition of a phoneme), and compared production errors between stimulation conditions. As expected, TBI02 exhibited an immediate and significant decrease in the total number of errors during motor thalamus stimulation (**Fig. 4f**). Importantly, the reduction was more evident for the tongue-twisters with a higher number of errors and specifically for /t/ and /k/ consonants (54.2% and 18.9% decrease for “Kick Tick” and “Tack Tad”, respectively), which were identified by the SLP as impaired in TBI02. We then quantified changes in the F1/F2 formants to assess articulatory control of vowels. Similar to TBI01, frequencies in both formants were significantly different with stimulation ON (decrease in the F1 formant frequencies for all tested vowels and increase in the F2 formant frequencies for /ae/ and /˄/), further suggesting an effect of the stimulation on mouth and tongue motor control.

Overall, our results demonstrate that motor thalamus stimulation yields a significant improvement in speech production in two subjects with mild-to-profound motor speech deficits. These compelling initial findings underscore the potential of this innovative approach as a groundbreaking treatment for dysarthria.

### Stimulation of the motor thalamus results in a clinically significant improvement in speech intelligibility after TBI

Finally, we sought to demonstrate whether the observed effects in respiratory, phonatory, resonatory, and articulatory control would result in a clinical improvement in speech intelligibility and quality. Because of the difference in their respective deficits, we personalized the assessment for the two TBI participants. For TBI01, we performed three clinical assessments with and without stimulation. We first had a blinded SLP administer the 2nd Edition Frenchay Dysarthria Assessment (FDA-2) to assess patterns of oral motor functions, with particular focus on the following subsections: Respiration, Lips, Palate, Laryngeal, and Tongue^47^. For each subsection, the SLP provided a score from A to E, representative of normal function to no function, respectively. Importantly, when scored on a 9-point scale with “E” corresponding to 1 and “A” corresponding to 9 across all subsections, the stimulation ON condition resulted in a 10-point improvement as compared to stimulation OFF (i.e., from 88 to 98/234, where 234 represents normal functions over all subsections, **Extended Data Figure 6**). Specifically, we found that motor thalamus stimulation immediately improved reflexes during swallowing (from severe to moderate impairments), paralleling the improvements observed in the videofluoroscopic swallow study (**Fig. 5a**). Additionally, stimulation improved lip control at rest and during an exaggerated smile (i.e., spread) from severe to mild impairments, suggesting that stimulation reduced facial paralysis and paralleled effects captured with the facial expression tasks. Finally, stimulation improved bilateral movements of the palate and elevation as reported by the SLP but slightly reduced palate maintenance (mild to moderate impairments). To assess speech quality, the same blinded SLP was provided with audio recordings of TBI01 performing speech tasks consisting of three repetitions of 2-word phrases, short (5 to 8-word) sentences^48^, and/or paragraphs with stimulation ON and OFF. For all four sessions evaluated, the SLP consistently rated the stimulation ON condition as being qualitatively more intelligible as compared to no stimulation (> 60% of the cases, **Fig. 5b**). We finally assessed speech intelligibility quantitatively with the single word section of the Assessment of Intelligibility of Dysarthric Speech test (AIDS)^49^: TBI01 was instructed to read aloud single word, and, listening to the audio, the same blinded SLP identified the word pronounced from a multiple choice of 12 words. Without stimulation, TBI01 presented with a profound speech deficit (<26% of speech intelligibility, i.e. 26% of words were correctly selected), which immediately improved when DBS was turned ON (8%, 20%, and 16% improvement for session 1, 2, and 3, respectively, **Fig. 5c**). Importantly, a 7% change in intelligibility can be considered a small clinically significant change; whereas a 15% change in intelligibility can be considered a large clinically significant improvement^50^.

**Fig. 5.**
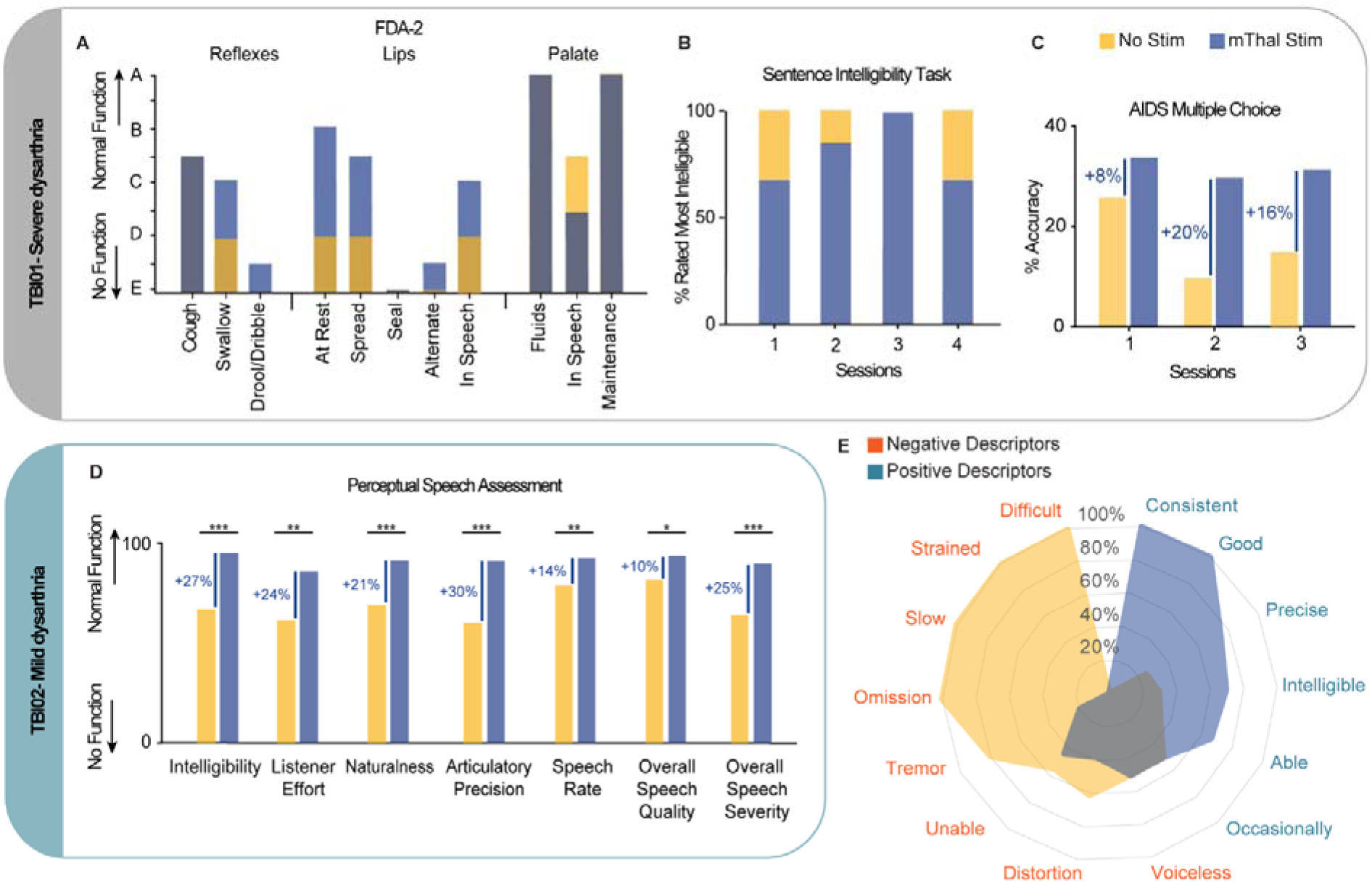
DBS results in a clinically relevant improvement in speech quality and intelligibility following TBI. Results are separated by participants to capture improvements in either profound dysarthria (TBI01, gray rectangle, panel a-c) or mild dysarthria (TBI02, teal rectangle, panel d). **(a)** Frenchay Dysarthria Assessment for the ON (blue) and OFF (yellow) stimulation conditions for TBI01 for the major items. See **Extended Data Fig. 6** for the remaining items. For each item, bars for the two conditions are overlapped. **(b)** Percentage of rated more intelligible speech for the 2-word phrases, short sentences, and paragraphs comparing stimulation ON (blue) and OFF (yellow) conditions for four sessions for TBI01. For each session, percentages were calculated over the total number of sentences tested for each condition. **(c)** Assessment of Intelligibility of Dysarthric Speech for stimulation ON and OFF conditions across three sessions for TBI01. Percentage accuracy represents how many words were correctly transcript by the SLP. **(d)** Perceptual speech measures rated by two blinded SLPs and five naive listeners for stimulation ON and OFF conditions for TBI02. Statistical significance assessed through two-tailed bootstrapping: p<0.05 (*), p<0.01 (**), p<0.001(***). **(e)** Polar plot of percentage for which a descriptor was used by the two SLPs to describe audio speech from TBI01 and TBI02. In orange negative descriptors (poor breath support, difficulty, missing, strained, slow, unable, distorted, voiceless, omissions, errors, wet) and in light blue positive descriptors (better breath support, consistent, good, precise, intelligible, able). In all panels, yellow indicates stimulation OFF and blue indicates stimulation ON.

Since TBI02 had nearly 100% speech intelligibility, we recorded TBI02 during spontaneous speech with and without stimulation. We then asked two certified SLPs and five naive native-English speakers blinded to the stimulation conditions to evaluate perceptual speech measures^51–54^ (see Methods) including: speech intelligibility, listener effort, speech naturalness, articulatory precision, speech rate, voice quality, and speech severity. Listeners reported a significant improvement across all metrics when the stimulation was ON (**Fig. 5d**) with a large clinically significant changes for intelligibility (+27%), effort (+24%), naturalness (+21%), articulatory precision (+30%), and speech severity (+25%).

Finally, we classified the frequencies of descriptive terminologies within the SLP evaluations of the sentences for TBI01 and the spontaneous speech for TBI02. These descriptors of speech quality were separated into two large categories: positive (e.g., ‘Precise’, ‘Concise’, ‘Intelligible’) and negative (e.g., ‘Distorted’, ‘Difficult’, ‘Slow’) descriptors. As expected, SLPs showed a preference for using more positive descriptors for sentences pronounced when the stimulation was ON, further supporting that DBS of the motor thalamus improves speech quality (**Fig. 5e**).

Overall, these results demonstrate that motor thalamus stimulation leads to clinically significant improvements in speech quality and intelligibility in two participants with a wide range of motor speech deficits. The observed enhancements suggest that this approach has substantial potential for restoring more natural speech in individuals with dysarthria.

## Discussion

In this study we report preliminary evidence that continuous DBS targeting the motor thalamus could immediately augment facial and oropharyngeal volitional motor output leading to clinically significant improvements in speech and swallowing functions in two participants who sustained partial lesions of the CBT.

At first our results seem to contradict years of literature. Indeed, subthalamic (STN) and thalamic DBS therapy are known to possibly produce adverse side effects that impact the articulatory and phonatory aspects of speech^55–58^. While the true extent of those secondary effects is difficult to gauge (as many studies rely on subjective reports, gross measures of speech production, and do not employ standardized testing), these effects might be attributable to the parameters commonly employed in clinical settings, particularly frequencies around 130 Hz, and site of stimulation. In fact, recent works that used objective evaluation of vocal parameters reported significant improvements in speech when lower frequencies (e.g., 60 Hz) were applied, as in our study^33–38^. Our choice of stimulation parameters (frequencies between 50-100 Hz) was driven by our recent understanding of mechanisms of action of motor thalamus stimulation on upper-limb function^18^: stimulation of the motor thalamus increases motor output by augmenting the recruitment of cortico-spinal and cortico-bulbar motor neurons, within the primary motor cortex, via excitatory synaptic inputs from the targeted thalamic nuclei. Importantly, because of the short duration of excitatory postsynaptic potentials on the membrane potential (5 to 20ms), synaptic-mediated excitatory inputs necessitate a stimulation frequency sufficiently high (> 40Hz)^59,60^ to induce sustained membrane depolarization, but sufficiently low (<= 100 Hz) to prevent presynaptic effects that reduce synaptic-mediated excitatory inputs^61–63^. Supported by the observed DCS-MEP potentiation in orofacial muscles, which aligns with our previous findings in primate models^18^, we hypothesize a similar mechanism for speech motor output. Further studies will now be necessary to confirm our hypothesis and to refine and validate the optimal stimulation parameters for enhancing speech motor function.

Regarding the site of stimulation, the dysarthric side effects of clinical DBS have been associated with the inadvertent spread of electrical currents to the CST and CBT tracts within the internal capsule^64^. In our study, we meticulously controlled for current spread by performing clinical evaluations of “capsular” motor side effects and by showing that thalamic stimulation neither increased muscle activation at rest nor induced any participant-reported discomfort or speech impairments. This absence of current spread to the capsule could be attributable to the low frequency stimulation we employed. Additionally, the motor thalamus includes three nuclei that have different preferential projection to the sensorimotor cortices: the ventral intermediate (VIM), which is the clinical target for thalamic DBS, the ventral oral posterior (VOP), and the ventral oral anterior (VOA) nucleus^65,66^. From an evidence-based yet speculative standpoint, the more anteriorly located nuclei (i.e., VOA and VOP) might represent a more optimal target for motor speech restoration. Indeed, our previous research on upper-limb motor function indicated that these nuclei, with their preferential projections to the primary motor cortex, are more efficacious in enhancing motor output^18^. This functional specificity may extend to speech-related motor functions, as the anterior motor thalamic nuclei not only project to the primary motor cortex but also connect with agranular cortical areas involved in speech motor production^67^, such as the premotor cortex and supplementary motor area^29–32^. This connectivity underscores the potential of targeted thalamic stimulation for restoring critical functions including speech and swallowing. However, the scope of our study was constrained by the clinical context in which participants were implanted, limiting our ability to assess the effects of stimulation on the different motor thalamus nuclei. Future research is imperative to comprehensively elucidate the optimal target of stimulation for the restoration of motor speech function.

DBS therapy in patients with PD or Essential Tremor (ET) provides immediate relief from movement disorder symptoms such as tremor, joint stiffness, and rigidity when the device is activated^25,68,69^. These immediate and robust improvements have been crucial for the acceptance of DBS technology for movement disorders. Therefore, achieving immediate improvements in speech and swallowing motor deficits upon activating DBS is essential for its clinical adoption in patients with cerebral lesions. This was the primary aim of our study: to assess the immediate benefits of thalamic stimulation. Although our small sample size limits conclusions about safety and efficacy, we report, to our knowledge, the first evidence of clinically significant improvements in dysarthria and dysphagia in patients with motor axon damage due to traumatic subcortical lesions. In this context, while recent clinical trials have shown that the surgical procedure for DBS implantation does not pose significant barriers to its clinical application for cerebral lesions^24,70^, to confirm efficacy, a larger longitudinal study is now needed to establish whether the observed immediate effects could support long-term recovery. Given prior research indicating enhanced outcomes when neuromodulation was complemented with speech therapy^71–73^, it is plausible that thalamic DBS, combined with targeted therapy, could provide a synergistic effect in treating dysarthria and dysphagia.

The most important limitation of our study is that results for speech and swallowing dysfunction are presented in only two TBI participants. However, effects of the stimulation on improving face motor cortex excitability and volitional motor control were reported in ten additional neurologically intact participants further suggesting the relevance of our approach. Moreover, despite the large differences between the two TBI participants in terms of age, severity of deficit, and time since the brain injury, both participants showed a significant improvement in speech quality and intelligibility with stimulation ON. Importantly, this amelioration was due to improved production of stop consonants and movement of the tongue, which are the aspects of speech typically more impaired in patients with dysarthria^1^. Additionally, effects of the stimulation were assessed with quantitative measures and were consistent over different tasks and clinical standardized tests further supporting the robustness of our findings. Finally, the consistency of our results across human subjects attests that the increased excitability of thalamic stimulation transcends etiological subtracts, further supporting the general applicability of this approach to a large spectrum of neurological conditions affecting volitional facial and oropharyngeal control. In this regard, it is important to highlight that TBI01 was the same participant of our previous work in which we demonstrated that motor thalamus DBS immediately improve arm and hand strength and volitional force control^18^, suggesting that our approach could be used to simultaneously treat speech and upper-limb deficits.

In conclusion, speech and swallowing control are a priority for millions of individuals with dysarthria and dysphagia, as underscored by recent advancements in neural engineering^74^. In this regard, several research teams have developed real-time brain-computer interfaces that detect intended speech from cortical neural signals and output it on a computer screen^75–80^, building on decades of brain decoding technologies for upper- and lower-limbs^74^. Inspired by these efforts, here we engineered a neurotechnology able to restore natural speech, i.e., a priority for patients and SLPs^81^, and improve swallowing functions. Given that DBS is an FDA-approved therapy for various motor disorders and the motor thalamus is a common clinical target for essential tremor treatment^25,26^, our approach holds promise as a novel therapeutic strategy for dysphagia and dysarthria.

## Methods

### Ethical considerations

All intra-operative procedures in human subjects were approved by the University of Pittsburgh Institutional Review Board (STUDY21040121). The behavioral experiments in human subjects were approved by the University of Pittsburgh Institutional Review Board (STUDY21100020 and STUDY2007011). All the recordings were carried out according to CARE guidelines and in compliance with the Declaration of Helsinki principles. The participants were informed of the procedure and they signed an informed consent, which included the consent for the use of all data collected during the experiment in scientific publications. Additionally, participants provided written consent to the use of photos, videos, and voice-recorded acquired during the experiments for research purposes and publications.

### Human participants

We recruited a total of 12 participants for this study (see **Supplementary Table 1** for clinical details and experiments performed for each participant). Specifically, we performed electrophysiological experiments on n=7 human subjects (S01-S07, 3 males and 4 females) of age 68.78 ± 3.43 (mean±std) who were undergoing DBS implantation of the motor thalamus to treat medically-intractable asymmetric Essential Tremor (ET) symptoms or of the subthalamic nucleus to treat Parkinson’s Disease (PD) symptoms (see **Supplementary Table 2** for clinical details). In all participants, we tested the least symptomatic side to better approximate normal function (right hemisphere was tested in 6/7 participants). We performed behavioral facial motor tasks in n=3 human subjects (2 male, 1 female) of age 29±6.94 who underwent surgical implant of stereo-electroencephalography (sEEG) for seizure monitoring and localization. Tests were performed during patient hospitalization in the epilepsy monitoring unit. Finally, we tested n = 2 participants who suffered a traumatic brain injury. TBI01 (male in his 40’s) had suffered severe traumatic brain injury resulting in diffuse axonal injury from a motor vehicle accident around 1 year before DBS implantation. The diffuse axonal injury involved the CST and was accompanied by bilateral edema of the cerebral peduncles and pons (**Extended Data Fig. 3a**, more pronounced in the left than the right hemisphere). Consequently, TBI01 suffered from hemiparesis of the right side and left upper extremity muscle weakness and tremor with decreased overall strength, balance, coordination, and endurance. According to SLP evaluations, the participant suffered severe motor speech impairment characterized by slow, uncoordinated, and imprecise speech with overall poor comprehensibility in unknown listeners’ context. The SLP also noted probable dysphonia with reduced vocal intensity, and hoarse vocal quality likely secondary to reported left vocal fold paralysis. With regard to swallowing function, the participant demonstrated oropharyngeal dysphagia characterized by suspected oral phase discoordination and possible airway compromise due to left vocal fold paralysis. The participant was recommended for bilateral DBS electrode implant to treat the post-traumatic tremor. TBI02 (male in his 50’s) sustained general right-side mild hemiparesis and mild dysarthria from a perinatal left hemispheric brain injury 40+ years prior to implantation of the motor thalamus electrode. The participant underwent sEEG implantation for seizure evaluation for treatment of drug-resistant epilepsy. Although the subject had received speech therapy immediately following the brain injury, SLP evaluation at the time of implant characterized the participant to have mild dysarthria of unspecific subtype. He demonstrated a reduced range of motion for both labial protrusion and retraction and distortion of phonemes worsening articulatory imprecision. Swallowing function in TBI02 was intact.

### Diffusion MRI and tractography analysis

Diffusion images for TBI01 and TBI02 were acquired on a SIEMENS Prisma Fit scanner using a diffusion sequence (2mm isotropic resolution, TE/TR= 99.2 ms/2490 ms, 257 diffusion sampling with maximum b-value 4010 s/mm²). The accuracy of b-table orientation was examined by comparing fiber orientations with those of a population-averaged template^82^. The tensor metrics were calculated using diffusion weighted images (DWI) with b-value lower than 1750 s/mm².

To quantify the white-matter fibers damage in TBI01 and TBI02, we used an advance diffusion MRI technique commonly used in patients with bilateral white-matter damage that allows for quantification of a decrease in anisotropic diffusion by applying a “tracking-the-difference” paradigm when compared to a normal population template: differential tractography^83^. Differential tractography was performed by comparing the high-definition fiber tractography (HDFT) of TBI01 and TBI02 against an age-and-sex-matched template constructed from a group of healthy controls of healthy quantitative anisotropy (QA). To capture neural injury, we highlighted tracks that displayed a 20% decrease in QA when compared to the template of healthy controls. We then quantified the composition of lesioned tracts based on a population-averaged atlas of human white matter tracts^82^ for the main fiber tracts: corticospinal tract, corticobulbar tract, and dentato-rubro-thalamic tract.

### Human intraoperative data acquisition and electrophysiology

Pairs of needle electrodes (Rhythmlink Columbia, SC) were placed subcutaneously in bilateral masseter (MASS), orbicularis oris (ORIS), mylohyoid (MYLO), and cricothyroid (CRICO) muscles to record EMG and evoked MEPs. At the beginning of the surgical procedure, a subdural strip electrode (6 contact platinum subdural electrode, AD-TECH Medical Instrument Corporation, Oak Creek, WI) was placed over the face representation of the primary motor cortex (M1) to perform direct cortical stimulation (DCS). The electrode position was optimized using results from DCS-MEPs of the contralateral face muscles. We selected the optimal subdural stimulation contact as the contact that elicited the largest MEPs in the masseter muscles and that produced a visual jaw contraction and tongue movement. DCS of the face representation of M1 was provided using trains of 5 stimulation pulses (0.5 ms) at 400 Hz every two seconds at stimulus intensities up to 15mA using an intraoperative neurophysiological monitoring system (XLTEK Protektor, Natus Medical, Ontario, Canada). Detailed information on the stimulation parameters used for DCS for each subject is reported in **Supplementary Table 2**. After implantation of the subdural strip electrode, the surgery continued following standard clinical procedures. Specifically, as part of the standard of care in our institution, DBS was performed using stereotactic techniques and with a robotic assisted device (ROSA, Zimmer-Biomed, Warsaw, IN, USA). The robot was used to insert three microelectrodes, while an Alpha Omega system (AO, Nazareth, Israel) was used for neurophysiological microelectrode recordings to map thalamic and subthalamic structures and identify the optimal location for implanting DBS electrodes. In ET patients undergoing DBS implantation of the motor thalamus, three different trajectories were used with the microelectrodes oriented in a rostral to caudal arrangement: (1) the anterior trajectory targeted the ventral oral anterior (VOA) nucleus (~10mm rostral to the posterior commissure – PC); (2) the center trajectory targeted the ventral oral posterior (VOP) nucleus (~8 mm rostral to PC); and (3) the posterior trajectory targeted the ventral intermediate (VIM) nucleus (~6 mm rostral to PC). For PD patients undergoing DBS implantation of the subthalamic nuclei, one trajectory was passed through the VOP nucleus (~8mm rostral to PC). For the electrophysiology experiments reported in this study, we stimulated from the macro contact of the microelectrode from the central trajectory in ET patients and from the microelectrodes passing through the VOP for PD patients. In both cases, the electrophysiology experiments were performed following completion of the clinical procedure, with the subject maintained under sedation with propofol. Specifically, DCS-MEPs were recorded by stimulating the optimal electrode contact over the face representation of the primary motor cortex, both without and with continuous stimulation of the motor thalamus (stimulation from the macro contact of the microelectrode) at 50, 80, and 100Hz with pulses of 100us and intensity of 4mA. Stimulation of the motor thalamus was delivered via the Alpha Omega system.

### Behavioral tests

Participants performed a series of behavioral tests depending on their impairments and time available. Behavioral tests were repeated over multiple sessions whenever possible (see **Supplementary Tables 1** and **4**). All behavioral tests were repeated without and with motor thalamus stimulation ON. For each session, amplitude of motor thalamus stimulation was selected to be the highest value that the patients could tolerate and that would produce a transient paresthesia in the facial muscles (see **Supplementary Table 3** for details on stimulation parameters for each participant). Tests were initiated only when the participants reported that the paresthesia disappeared. For all tasks, a computer monitor was placed in front of the participants, about 25-30 inches away from their face to instruct them on the tasks. Participants were instructed to initiate the trial after a cue that paired a short tonal sound with the appearance of the action prompt on the screen. For tasks with time constraints, participants received feedback of the remaining time during the trial and visual feedback to stop the instructed action after the allotted time. A directional microphone was placed in front of the participant to record both the participants’ speech and the auditory go cue from the monitor without occluding the behavioral task displayed on the monitor. TBI01 completed all behavioral tasks sitting in a chair while subjects TBI02 and S08-S10 completed their tasks sitting in an upright posture in standard hospital beds. Stimulation ON and OFF conditions were randomized across subjects for S08-S10 and across sessions for TBI01 and TBI02.

#### Facial tasks

To evaluate stimulation effect on voluntary facial movement, four facial tasks were designed to evaluate the range of movement and aperture of facial articulators critical to speech and swallowing: 1) protruding tongue in/out (TT); 2) smiling (TS); 3) opening/closing mouth (TO); 4) puckering the lips (TL). For all tasks, each participant was instructed to alternate between a neutral facial expression and the designated facial task as fast and as large as they could within 15 seconds. The study was repeated with and without motor thalamus stimulation. Video of the facial movements and surface EMG of the masseter, orbicularis oris, and mylohyoid muscles were recorded across all tasks. All subjects completed at least one session of the facial tasks. However, video quality was too poor for kinematic tracking for TBI02 and so data were not included in the analysis. S08, S09, and S10 only performed these tasks within a single session whereas TBI01 participated across multiple sessions. Only sessions where TBI01 could complete the tasks for all stimulation conditions while sitting in a consistent upright posture were included (see **Supplementary Table 4**).

#### TBI01 - single words

To measure effects of stimulation on the four subsystems of speech, TBI01 was tasked to repeat single words five times as clearly as possible. We tested words composed of one to three syllables (n = 37 one syllable, n = 20 two syllables, and n = 2 three syllables) and ensured that all American English phonemes were tested in at least one of the eight experimental sessions. See **Extended Data Fig. 5** for the full list of tested words and their phoneme composition. TBI01 performed this task across multiple sessions. Only sessions in which TBI01 could complete all tasks for all stimulation conditions were included in the data analysis (see **Supplementary Table 4**).

#### TBI01 – sentences

To measure effects of stimulation on speech quality, TBI01 was tasked to repeat aloud two-word phrases, short (5 to 8-word) sentences, and/or paragraphs for a set number of repetitions. For the two-word phrases, we were looking to highlight sound differences in monosyllable minimal pair words (e.g. *beep-peep, tip-dip*), by having TBI01 repeat the words in the phrase “To *X*” five times. This phrase construction ensured that the consonant sound contrasts always followed a vowel rather than being in the word-initial position. For the short sentences, five short sentences were selected from the 460 sentences in the MOCHA-TIMIT database^48^ and TBI01 was instructed to repeat each sentence three to four times. We deployed the MOCHA-TIMIT database because sentences are phonetically balanced and include the main connected speech processes for American English. Finally, TBI01 performed single recitations of classic speech therapy reading paragraphs including the Grandfather passage^84^, the Caterpillar passage ^85^, and the Elicitation paragraph^86^. These paragraphs are phoneme balanced, include high-frequency words, vary sentence length throughout the paragraph, and contain most clusters of standard American English. TBI01 performed these tasks across multiple sessions, while audio recordings were acquired. Only sessions in which the participant could complete all tasks for all stimulation conditions were included in the analysis (**Supplementary Table 4**).

#### TBI02 - tongue twisters

In order to challenge the milder speech deficits observed in TBI02, we had the participant repeat 2-word tongue-twisters as fast as possible for 20 seconds^42^. The tongue-twister pairs were composed of words that were monosyllabic, following a consonant-vowel-consonant structure (CVC). These word pairs would have either a consonant mismatch (C_a-V-C_b vs C_c-V-C_b), a vowel mismatch (C_a-V_a-C_b vs C_a-V_b-C_b), or both (C_a-V_a-C_b vs C_c-V_b-C_b). We primarily selected words that matched words tested also in TBI01 (**Extended Data Fig. 5**). TBI02 was tested over 2 different sessions (**Supplementary Table 4**).

#### Videofluoroscopic swallow study

Since only TBI01 self-reported swallowing deficits, videofluoroscopic swallow study was performed exclusively with this participant. At the time of the test, TBI01 was on a regular diet with mildly thick liquids. Adhering to these reports, the videofluoroscopic swallow study^41^ (i.e., modified barium swallow, MBS) consisted of challenging the participant with the following adjusted consistencies: 1) mildly thick liquid via straw, 5ml bolus; 2) mildly thick liquid via straw, 10ml bolus; 3) unmeasured free straw sip; 4) pudding. By standard protocol, trials consisting of swallowing varying volumes of mildly thick liquid (5ml, 10ml, and unmeasured free straw sip) were repeated twice. However, one trial of the unmeasured free straw sip was excluded from analysis due to failure to capture all complete phases of swallowing. Videofluoroscopy was acquired for each swallow at a frequency of 30 frames per second. The study was repeated with and without motor thalamus stimulation at 55 Hz.

### Electromyographic recordings and analysis for intraoperative muscle activity

MEP trials were recorded for 100 ms following each DCS stimulation burst at a sampling frequency of 6000 Hz using the XLTEK system. To quantify DCS-MEPs recorded from triggered electromyographic activity, we computed the stimulation triggered averages (EMG from 10 ms to 75 ms after stimulation) and calculated the area-under-the-curve (AUC) for each muscle.

### Electromyographic recordings and analysis during behavioral tasks

To quantify stimulation effect on muscle activation during behavioral facial tasks, we collected surface EMG data from subjects S08 and S10. For S08, we applied disposable surface EMG electrodes from Rhythmlink on the facial muscles (masseter, orbicularis oris, mylohyoid) contralateral to the stimulation hemisphere and recorded activity using the Ripple Neuro Grapevine and Trellis software at a sampling frequency of 1000 Hz. We collected bilateral facial muscles (masseter, orbicularis oris, mylohyoid) from S10 using a wireless EMG system (Trigno, Delsys Inc) at a sampling frequency of 8 kHz. For both participants we used the same data processing pipeline where raw EMG traces were filtered using a second-order butterworth filter: first bandpass filter between 60 to 500Hz, then low-pass filter at threshold of 6Hz. For each movement repetition, movement initiation and completion were identified manually from visual inspection of the kinematic traces and videos. AUC was calculated for all movement repetitions for each task for each participant both during the movements and during neutral expressions.

### Kinematic recordings and analysis

Kinematics of facial tasks for participants S08-S10 and TBI01 were recorded by a GoPro® Camera and analyzed through DeepLabCut, a neural network for 2D and 3D markerless pose estimation based on transfer learning^87^. All videos recorded of participants performing facial tasks were initially processed using Adobe Premiere Pro to standardize framing and lighting across stimulation conditions. Kinematic facial articulators were manually labeled for a minimum of 40 frames per video clip. The labeled facial structures included jaw, cheek, mouth, upper lip, lower lip, nose, and chin. We employed DeepLabCut and a cloud computing cluster (Google Colab Pro) to train a neural network and extrapolate the articulator positions across all frames. Each dataset for each participant was trained for a minimum of 300,000 iterations. Visual inspection from LT and JH confirmed facial articulators automatically labeled by trained DeepLabCut models. Final x- and y-coordinates of each articulator across all tasks were used for kinematic analysis. For each movement repetition, movement initiation and completion were identified manually from visual inspection of the kinematic traces and videos. For each facial articulator, we calculated amplitude and velocity of movement. Additionally, we quantified mouth aperture by calculating the area between three (for videos capturing side profile) or four (for videos capturing front profile) representative facial articulators.

### Audio Recordings and analysis

All voice recordings were collected using a Sennheiser MKE 600 microphone (Sennheiser electronic SE & Co. KG, Germany) that was then amplified through a M-track Duo (48 kHz, 2-channel USB audio interface with 2 Combo Inputs, Crystal Preamps, and Phantom Power, M-audio, Rhode Island, USA) and finally digitally processed, and recorded using the Ripple Neuro Grapevine and Trellis software at a sampling frequency of 30 kHz. Each audio file was then cut in single word using a semi-automatic MATLAB script that removed pauses and non-task relevant sounds. The cutting process was verified by both auditory and visual inspection before the cut words were saved as both MATLAB structures and WAV files. To determine the beginning and end of individual phonemes, we used the Montreal Forced Alginer (MFA) to train an acoustic model to align phoneme transcripts to audio files. Specifically, we used the CMU ARPABET library to train the model as it is a dictionary for North American English^88^. Models were trained individually for each participant (TBI01 and TBI02) including all sessions for each individual. The final output of the model was grapheme “TextGrids” compatible with PRAAT^89^. Phoneme alignment was then inspected and confirmed for each WAV file using a custom PRAAT script^89^. The final grapheme information was added to the MATLAB structures, storing both single words and their individual phoneme traces. All MATLAB analysis used these structures and all PRAAT^89^ analysis used the WAV and TextGrid datasets.

### Speech Assessment

Due to the profound deficits of TBI01, we always excluded the first trial of each single word. This was to reduce any confounding/misleading effects from initiation variability and errors, including coughing, speaking over the tonal go cue, poor breath support, and whispering the word. Each word within a session was considered as a single word, resulting in a dataset of 108 words for TBI01. We included all speech trials for TBI02 and each word within a session for a total of 14 words. We calculated several speech metrics to assess effects of the stimulation on speech productions.

#### Speech Intensity

To assess changes in speech intensity with motor thalamus stimulation, we first decomposed the audio signal of every recorded word into a spectrogram. The spectrogram was calculated every 5 ms using a 25 ms Hanning window and the frequency range was limited to 50-10000 Hz. We calculated the total power in decibels for each time step and then applied a second-order 1D median filter to the trial trace. For each trial we calculated speech intensity as the median value of the smoothed trace.

#### Voice Breaks

To evaluate the effects of the stimulation on respiratory control, we determined voice breaks for each word individually using PRAAT’s voice report function^89^. We first calculated a pitch object using a filtered autocorrelation method (recommended for voice pathology) over the entire sound bit with the standard settings save for pitch range which we adjusted to 75-400 Hz and the voice threshold which we reduced to 0.25 to better evaluate irregularities in the speech signal. We then created a PointProcess object to define the pulse train for the signal before running PRAAT’s voice report function^89^ with its standard settings. Voice breaks were defined as any inter-pulse intervals between voiced frames that lasted longer than 16.67ms.

#### Cepstral Peak Prominence-smoothed

In order to assess the effects of the stimulation on phonatory control, we computed the Cepstral Peak Prominence-smoothed (CPPS) for each unique trial of each word, reflecting the harmonic structure of the speech signal and the amplitude of the cepstral peak^90,91^. We employed a PRAAT script to calculate the power-cepstrogram from each word’s spectrogram, and subsequently derived the CPPS using the “Get CPPS” function with default settings from Hillenbrand et al., 1996^44^. The CPPS values from all words and sessions were concatenated to assess the percent change in voice quality between stimulation OFF and stimulation ON conditions.

#### Fatigue

To determine whether stimulation affects the endurance aspect of speech, we timed the duration TBI01 spent in completing the single-word tasks as a measurement of fatigue. Video recording from sessions where the order of words tested remained consistent across stimulation conditions were analyzed. Time duration necessary for TBI01 to complete five repetitions of the first and last five unique words tested between stimulation ON and OFF was measured by time stamp from video footage. The start time was marked by the auditory go cue and end time was identified by the termination of the last repetition of the last word. Longer time duration suggested increased fatigue from the speech task.

#### Consonant Articulation

To determine whether there was improvement in the articulation of consonants, for TBI01 we perceptually assessed the presence or absence of each phoneme. Specifically, a researcher (EG) blinded both to the stimulation condition and session listened to the word while being presented with a transcript of the word and its phonemes. The researcher then classified each phoneme as being 1) *present*, or 2) *distorted* or *fully absent*. The trial’s recording was replayed as many times as necessary until all phonemes in a given word were characterized. Next, we calculated the percentage of phoneme presence for stimulation OFF and stimulation ON for each session by concatenating the phonemes across words. For TBI02, instead, we identified four common production errors for the tongue-twisters: pronunciation of incorrect phoneme (e.g. /tæk/ instead of /tɪk/), speech arrest, slurring words, and stutter words. Specifically, speech arrest was identified as struggling to initiate or complete a word. Slurring was identified when the listener would identify consonants that sounded messy, prolonged, or unclear. Stuttering was identified when there was repetition of a phoneme prior to completing the word. The identification of these errors was made separately by three (EG, IM, SN) native English speakers who were blinded to the stimulation conditions. To reduce bias, the listeners had to record both the timing and type of error that they observed. An error was only counted if at least two of the listeners had identified the exact same timing and type of error.

#### Formant Frequencies

To determine whether there was improvement in the articulation of vowels, we quantified changes in the formant frequencies of formant F1 and F2. We estimated formant frequencies using linear predictive coding (LPC) coefficients. To build the LPC model, we first preprocessed the audio data. This involved extracting individual vowel phoneme segments from the recorded words and applying an all-pole pre-emphasis filter *P*(*z*) = 1 – 0.63*z*^-1^ to enhance higher frequency components and improve the signal-to-noise ratio. Next, we created a spectrogram using a Hamming window, segmenting the data into 15 ms windows with 80% overlap. We then calculated the LPC coefficients, using a model order determined by *F_s_*/1000 + 2 (F_s/1000 + 2), where *F_s_* is the sampling frequency. We identified the first four formants (F1, F2, F3, F4) as the first four roots of the LPC spectrum that had frequencies greater than 90 Hz and bandwidths less than 400 Hz. All formant frequency calculations were performed in MATLAB. We verified the results by comparing them with formants identified using PRAAT’s default settings, confirming consistency and accuracy. Given that F1 and F2 correspond to vowel openness and tongue position^46^, respectively, we next assessed how these values change with stimulation. For each session, we identified all unique vowels tested to determine the average F1 and F2 values for each trial. Since formant values are calculated in 15 ms intervals over the duration of the sound, each trial of a phoneme produces multiple formant values. This is important because different words may require different durations for the same vowel, and voice breaks during the trial can lead to inaccurate formant estimates. To address these issues and minimize bias in the formant range, we preprocessed the formant data by smoothing and time-warping it to ensure consistency across trials. We smoothed the first two formant signals using a first-order median filter. For time-warping, we applied spline interpolation to resample the data to 100 points for the entire vowel. We then truncated the data to cover 70% around the center of the vowel (i.e., points 15 to 85), ensuring that only the most stable portion of the vowel sound was used in calculating the averages for each trial. The magnitude of change was calculated as the percent change from the average formant value in the stimulation OFF condition to the ON condition.

### Perceptual Speech Assessment

Prior to this assessment, all audio recordings were edited in Adobe Premiere Pro to blind stimulation conditions. Because of the profound dysarthria, perceptual speech assessment of 2-word phrases, short (5 to 8-word) sentences^48^, and/or paragraphs from TBI01 was performed only by a trained SLP blinded to the experimental design and simulation conditions (ST). The designated phrases, sentences, or paragraph for each task was presented to the SLP within the video at the beginning of each task. Instead, for TBI02, we employed a validated auditory-perceptual assessment of dysarthria through Qualtrics to trained (n=2) and naive listener (n=5) groups blinded to stimulation conditions. In this case, the transcript of the speech was not provided to the listener. Each listener rated audio samples from TBI02 using separate visual analog scales (VASs) of 0-100 across 7 features, with 0 indicating “severe speech impairment” and 100 as “normal speech, no impairment”, respectively. Description and evaluation instructions for each feature is listed below:

1. **Speech Intelligibility:** Judge how well the speech is understood.
2. **Listener Effort**^51,52^: How effortful was it for you to understand this speaker?
3. **Speech Naturalness**^54^: Rate the naturalness of speech

a. Pay attention to how the speech compares with that of non-disordered speech as well as if it conforms to standards of rate, rhythm, intonation, and stress patterning and if it conforms to the grammatical structure of the utterance being produced.
4. **Articulatory Precision**: Judge how precise the speech is; scale severity based on whether some sounds vs. most sounds were slurred.
5. **Speech Rate**: Rate severity based on whether speakers are somewhat slow vs. very slow.
6. **Overall Voice Quality**: Rate severity based on how the voice compares with that of non-disordered voice (e.g., breathiness, roughness, harshness, etc.)
7. **Overall Speech Severity**^53^: Rate the overall severity of speech paying attention to the following:

a. Voice (quality – breathy, noisy, gurgly, high pitch, too low pitch, or sounds normal)
b. Resonance (too nasal, not nasal in the right places, sounds like they have a cold, or sound normal)
c. Articulatory precision (some sounds are crisp or slurred or somewhere in between or sounds normal)
d. Speech rhythm (the timing of speech doesn’t sound right or sounds normal)
e. Pay attention to overall speech naturalness and prosody (melody and timing of speech). Do not focus on the speaker’s intelligibility or how understandable each sentence is.

In addition to the auditory-perceptual assessment, the SLP clinical assessments always include descriptive impressions evaluating aspects of speech that were not directly quantifiable. Nevertheless, these descriptions convey critical qualitative insights to speech function. Therefore, to evaluate the effect of stimulation on overall clinical speech impression for TBI01 and TBI02, all descriptive reports conducted by a trained SLP (ST) for both participants were aggregated and descriptors were classified as positive (Consistent, Good, Precise, Intelligible, Able, Occasionally) and negative (Difficult, Strained, Slow, Omission, Tremor, Unable, Distortion, Voiceless). For TBI01, the reports analyzed consisted of SLP’s descriptive evaluation from the sentence intelligibility tasks. For TBI02, the report analyzed consisted of the Speech Initial Evaluation. Only descriptors consistently mentioned across both stimulation conditions at least once were included within the analysis.

### Analysis of videofluoroscopic swallow study

In addition to the SLP evaluation of the videos, we performed quantitative analysis of the swallow duration and amount of residue. Specifically, swallow duration was calculated by number of frames of radiology video report from first observation of modified barium within the oral cavity to the end of the swallow for all trials. Behavioral signs indicative of swallowing completion included clearance of barium through the upper esophageal sphincter and relaxation of the sphincter to its resting state. Area of Residue was quantified through ImageJ. Specifically, one representative frame capture for each phase-matched trial with the maximal visible modified barium bolus was manually labeled as region of interest (ROI) from the videofluoroscopic swallow radiology video report. Total area of ROIs was calculated in Pixel2 then normalized to the pixel area of the entire frame capture.

### Statistical Procedures

All statistical comparisons of means presented in this manuscript were performed using the bootstrap method, a non-parametric approach which makes no distributional assumptions on the observed data. Instead, bootstrapping uses resampling to construct empirical confidence intervals for quantities of interest. For each comparison, we construct bootstrap samples by drawing a sample with replacement from observed measurements, while preserving the number of measurements in each condition. We construct 10,000 bootstrap samples and, for each, calculate the difference in means of the resampled data. We employed two tailed bootstrapping with alphas of 0.05 (95% confidence interval), 0.01 (99% confidence interval), or 0.001 (99.9% confidence interval). The null hypothesis of no difference in the mean was rejected if 0 was not included in the confidence interval of the corresponding alpha value. If more than one comparison was being performed at once, we used a Bonferroni correction by dividing the alpha value by the number of pairwise comparisons being performed. Nonparametric paired t-tests were used specifically for the analysis of duration, residue area of swallowing function, and fatigue of speech function. For videofluoroscopic swallowing study, due to the varying volume and consistency of content swallowed across each trial (5 ml, 10 ml, free sip of liquid, and pudding), swallowing time and residue also varies for each trial. Therefore, comparison between stimulation conditions ON/OFF was paired across trials where the same content was swallowed. Similarly, fatigue analysis evaluates the time duration spent repeating the first and last five words tested within each session. Since the length of the words tested for each session were distinct, the time necessary to complete repetitions of the first and last five words varies. Hence, to evaluate stimulation effect on speech fatigue, duration was paired within each session where the words tested were consistent between stimulation ON/OFF conditions.

### Reporting Summary

Further information on research design is available in the Nature Research Reporting Summary linked to this article.

## Data availability

The main data supporting the results in this study are available within the paper and its Supplementary Information. All data generated in this study and software will be uploaded in a public repository upon acceptance of the manuscript. Raw data will be available upon reasonable request to the corresponding author.

## Data Availability

All data produced in the present study are available upon reasonable request to the authors.

## Acknowledgements

We thank Isabella Bushko for the design of fig. elements and Dr. Anna Chrabaszcz for discussion about the selection of speech motor tasks for TBI01. The study was executed through the support of internal funding from the Department of Physical Medicine and Rehabilitation at the University of Pittsburgh to EP and from the Department of Neurological Surgery at the University of Pittsburgh to JGM. Additionally, JGM discloses support for the research described in this study from the Hamot Health foundation and the National Institute of Health (R01NS122927). BZM was supported by R01EY028535 and R01NS089069.

## Author contributions

EP and JGM conceived the study and secured funding. EG, LT, AD, JH, BM, JF, DC, JGM, and EP designed the experiments. EG, LT, AD and JH collected all data with the assistance of TC, JGM, and EP. EG, LT and AD analyzed all the data with assistance from JH for the processing of the DTI data and the deep lab cut data for the kinematic analysis and from IMM and SN for the speech data. EP, JGM, DC, JF, and KLS helped with data interpretation. ST is a trained SLP and performed all clinical assessments. KLS (trained SLP) helped with assessments of audio recordings. TC and JGM implemented patient recruitment, eligibility and monitoring and coordinated study management. KF helped with patient scheduling. JGM performed all the surgeries. DC performed the subcortical mapping of motor thalamus for the DBS implantations. GMA and DC collected the electrophysiology human data. EP, JGM, EG and LT wrote the paper with assistance from DC and KLS and all authors contributed to its editing.

## Competing interests

BZM holds intellectual property (PCT/US2019/064015) associated with predictive analytics in neuromedicine, and is co-founder and CSO of MindTrace Technologies, Inc.

## Additional information

### Extended Data Figures

**Extended Data Figure 1.**
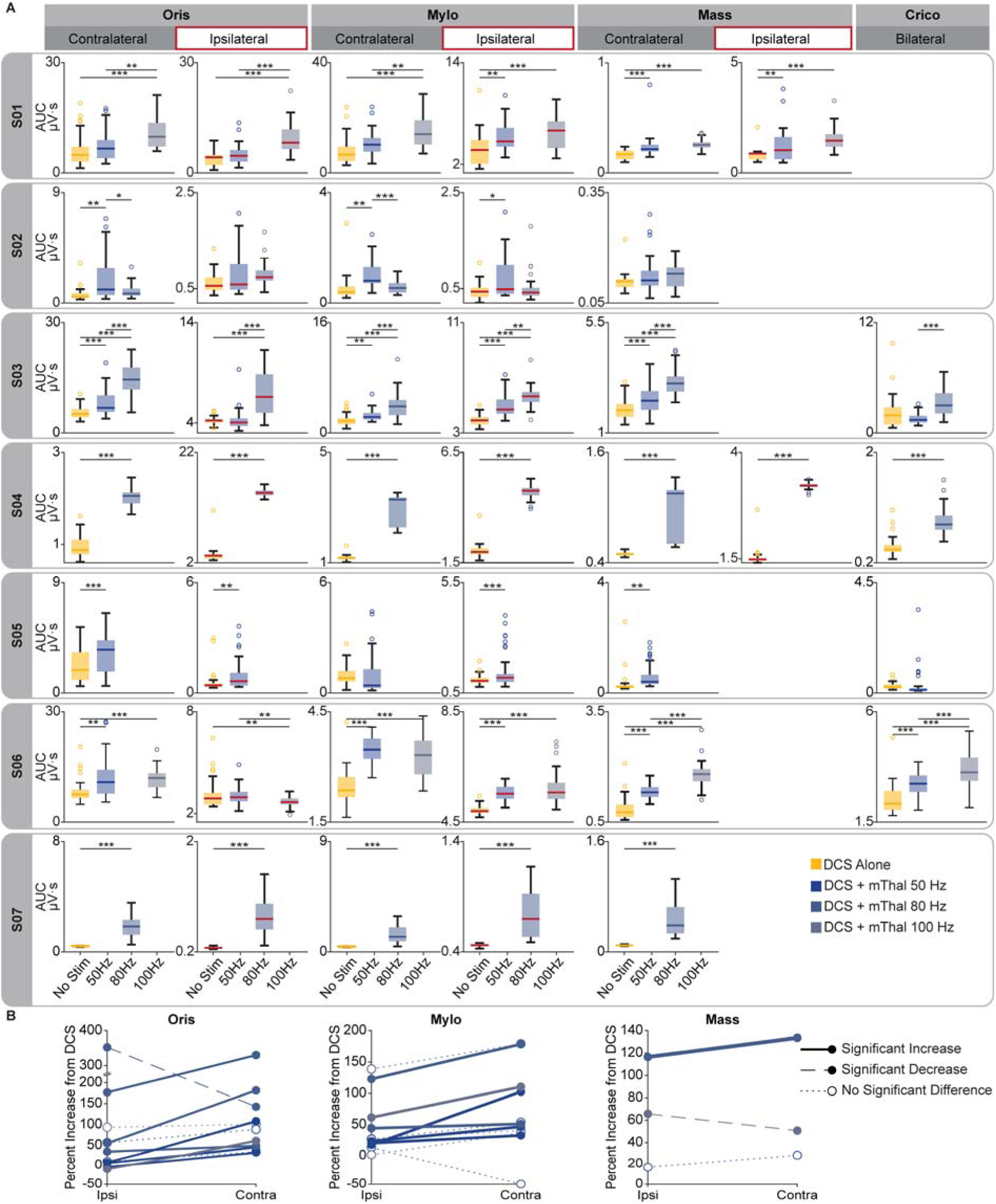
Stimulation of the motor thalamus potentiates MEPs of contralateral and ipsilateral orofacial muscles. **(a)** Boxplots of MEPs AUC amplitudes of different muscles (either contralateral, i.e., on the other side of the face from DCS and motor thalamus stimulation, or ipsilateral, i.e., on the same side of the face, or bilateral) with DCS alone and DCS paired with motor thalamus stimulation at 50, and/or 80 Hz, and/or 100 Hz. All subjects (S01-S07) are reported. Repetitions per condition for each patient: S01 n=30, S02 n =31, S03 n = 31, S04 n = 35, S05 n = 46, S06 n = 31, S07 n = 41. Mass: masseter, Oris: orbicularis oris, Mylo:, mylohyoid, Crico: cricothyroid. For all boxplots, the whiskers extend to the maximum spread excluding outliers. Central, top, and bottom lines represent median, 25^th^, and 75^th^ percentile, respectively. **(b)** Comparison of the percent increase of AUC for motor thalamus stimulation ON as compared to no stimulation for ipsilateral and contralateral pairs of muscles for all patients pulled together. Lines are colored by motor thalamus stimulation intensity (i.e., 50, 80, or 100Hz). Bolded lines represent MEPs for which contralateral potentiation was greater than ipsilateral potentiation (p<0.05); dashed lines represent MEPs for which ipsilateral potentiation was greater than contralateral (p<0.05); and dotted lines represent no significant difference between contralateral and ipsilateral muscles. For all panels, statistical significance was assessed with two-tail bootstrapping with Bonferroni correction: p<0.05 (*), p<0.01 (**), p<0.001(***). Artifact contamination from DCS in the ipsilateral masseter muscle prevented data collection in several cases. The cricothyroid muscle was only assessed after data collection for the first two study participants to increase the ability to quantify potential speech improvements physiologically.

**Extended Data Figure 2.**
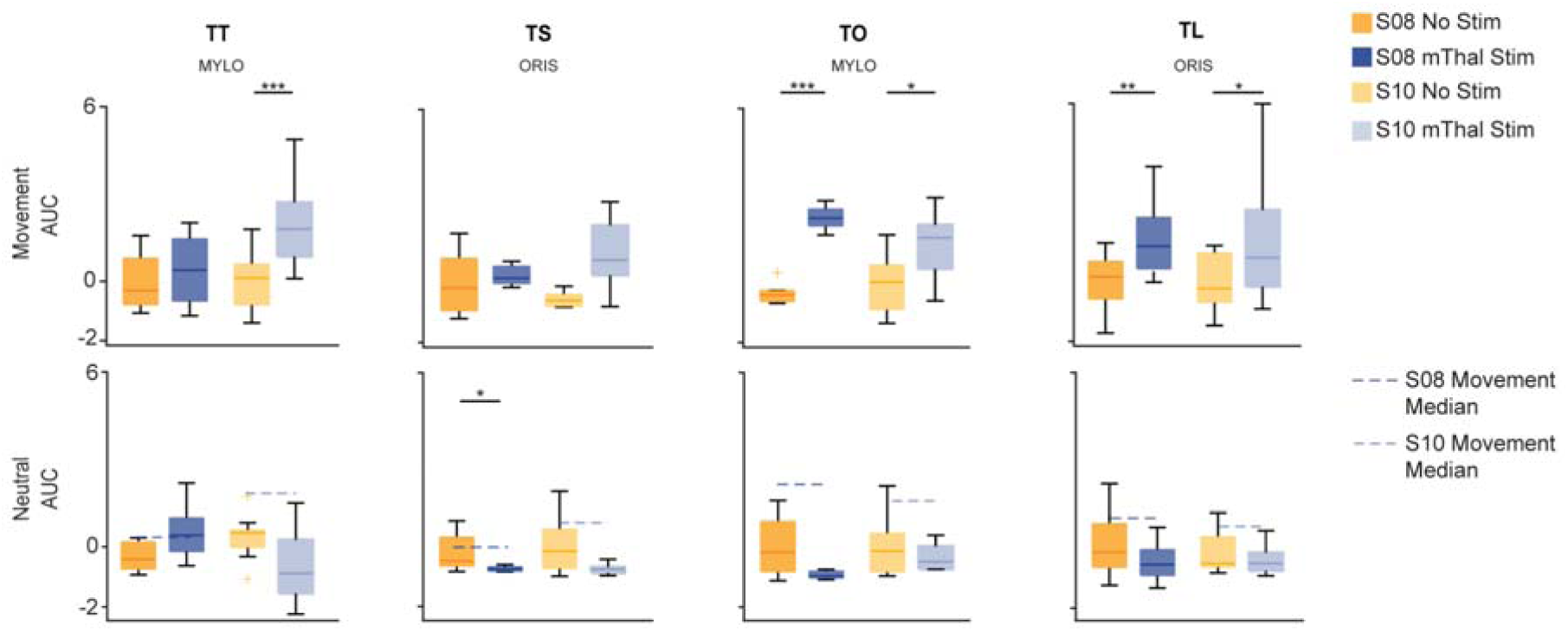
EMG activation during facial tasks and at rest. *Top*: Boxplots of z-score of EMG AUC for mylohyoid and orbicularis oris during movement phase of facial tasks (n=2) without (yellow) and with (blue) motor thalamus stimulation. This is the same plot of **Figure 2c**. *Bottom:* Boxplots of z-score of EMG AUC for mylohyoid and orbicularis oris during neutral phase of facial tasks. Dotted line represents median AUC from movement phase of respective facial tasks with stimulation.

**Extended Data Figure 3.**
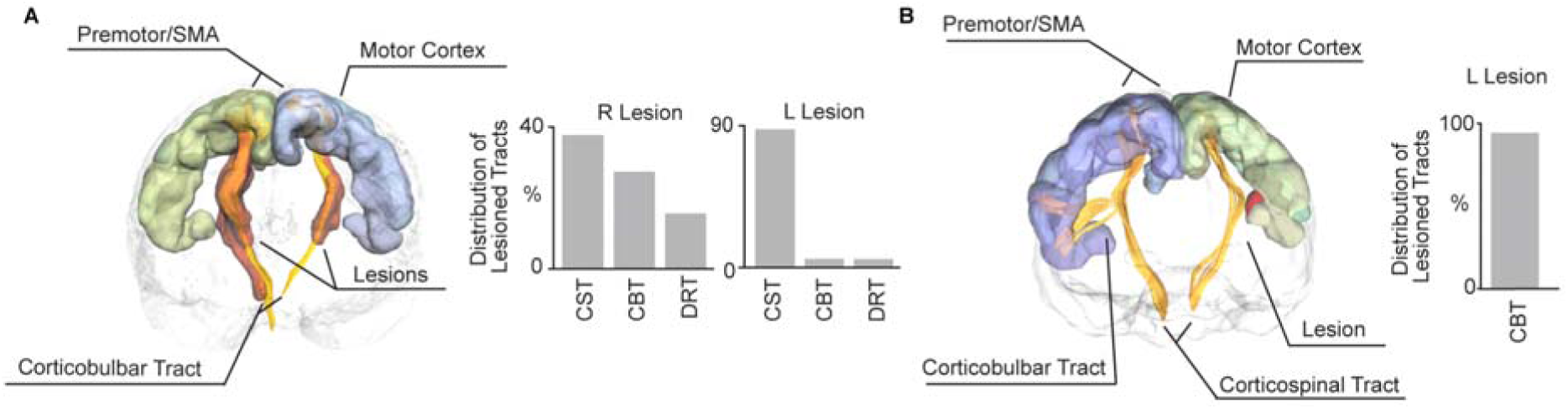
Lesioned white-matter fibers for TBI01 and TBI02. **(a)** *Left*: Tractography with lesion highlighted in red for TBI01. Right: Amount of lesioned tracts for both hemispheres (CST: corticospinal tract, CBT: corticobulbar tract, DRT: dentatorubrothalamic tract). This panel was adjusted from Ho & Grigsby et al., 2024^18^. **(b)** *Left*: Tractography with lesion highlighted in red for TBI02. *Right:* Amount of lesioned tracts in the left hemisphere (unilateral damage).

**Extended Data Figure 4.**
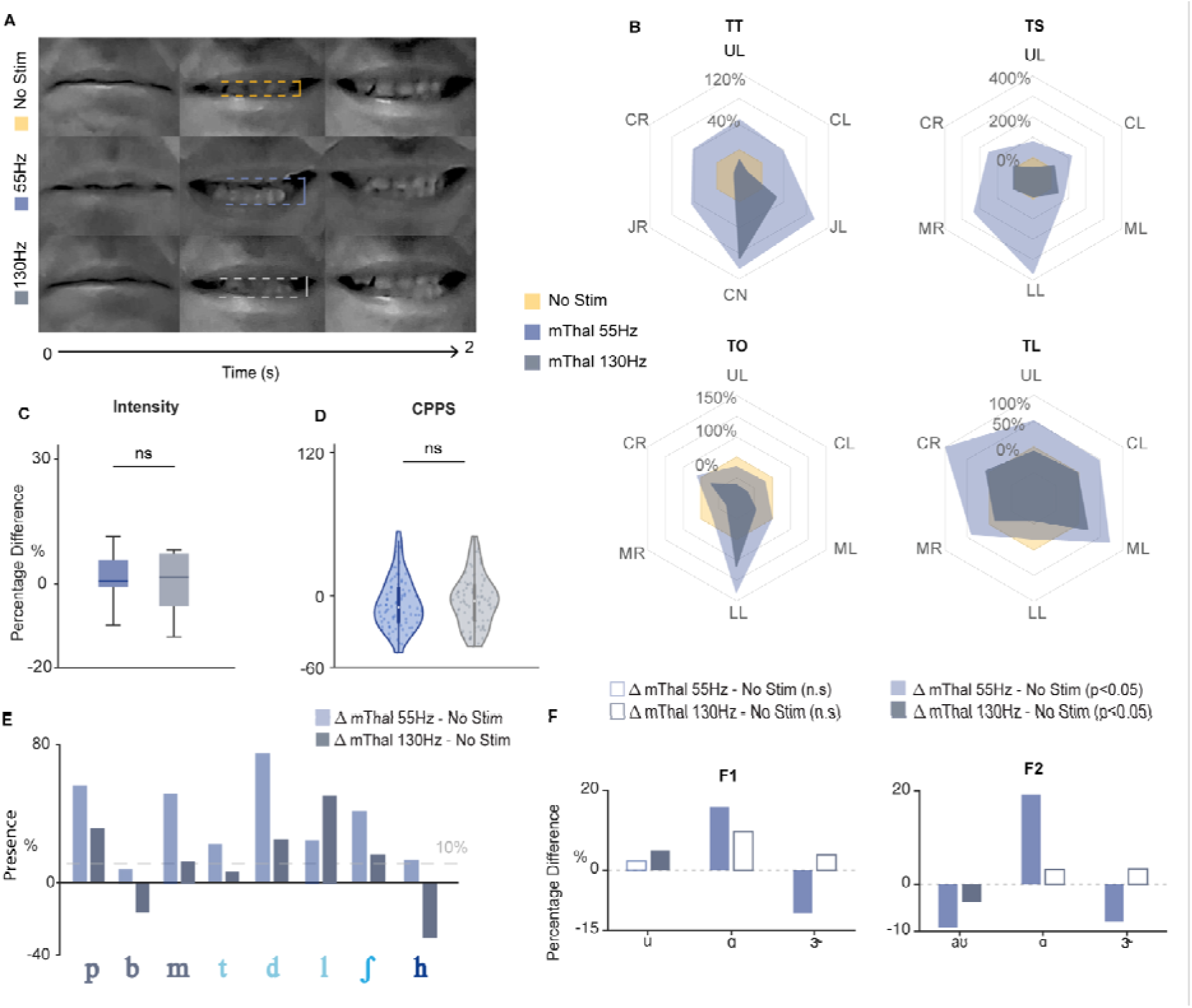
Comparison of effects for motor thalamus DBS at 55 and 130 Hz for TBI01. **(a)** Frame captures from video showing TBI01 maximum range of motion of the lips during smile task for no stimulation (yellow), motor thalamus (mThal) stimulation at 55 Hz (blue), and motor thalamus stimulation at 130 Hz (grey). **(b)** Radial plots of normalized change in amplitude of facial articulators during the four facial tasks for motor thalamus stimulation at 55Hz (blue) and motor thalamus stimulation at 130 Hz (gray). Facial articulators include upper lip (UL), jaw left (JL), jaw right (JR), cheek left (CL), cheek right (CR), mouth left (ML), mouth right (MR), chin (CN), and lower lip (LL). **(c)** Boxplots of speech intensity for stimulation ON at 55 Hz (light blue) and stimulation ON at 130 Hz (gray) conditions for single words. **(d)** Violin plot of CPPS for motor thalamus stimulation at 55 Hz and 130 Hz for single words. Each dot (n = 22) represents the trial average of a single word from a single session. For panels c and d, statistical significance was assessed with two-tailed bootstrapping with Bonferroni correction. No significant difference was found between the two conditions. **(e)** Percentage of perceptual presence for consonants tested when stimulation was ON at 55 Hz or 130 Hz, as compared to stimulation OFF. All bars are normalized to their stimulation OFF average. **(f)** Percentage of change in F1 and F2 formant frequencies between stimulation OFF and stimulation ON at 55Hz or 130Hz, respectively. Filled bars correspond to statistically significant changes (p<0.05), while empty bars correspond to non-significant changes. Statistical significance was assessed with two-tailed bootstrapping with Bonferroni correction. All bars are normalized to their stimulation OFF average.

**Extended Data Figure 5.**
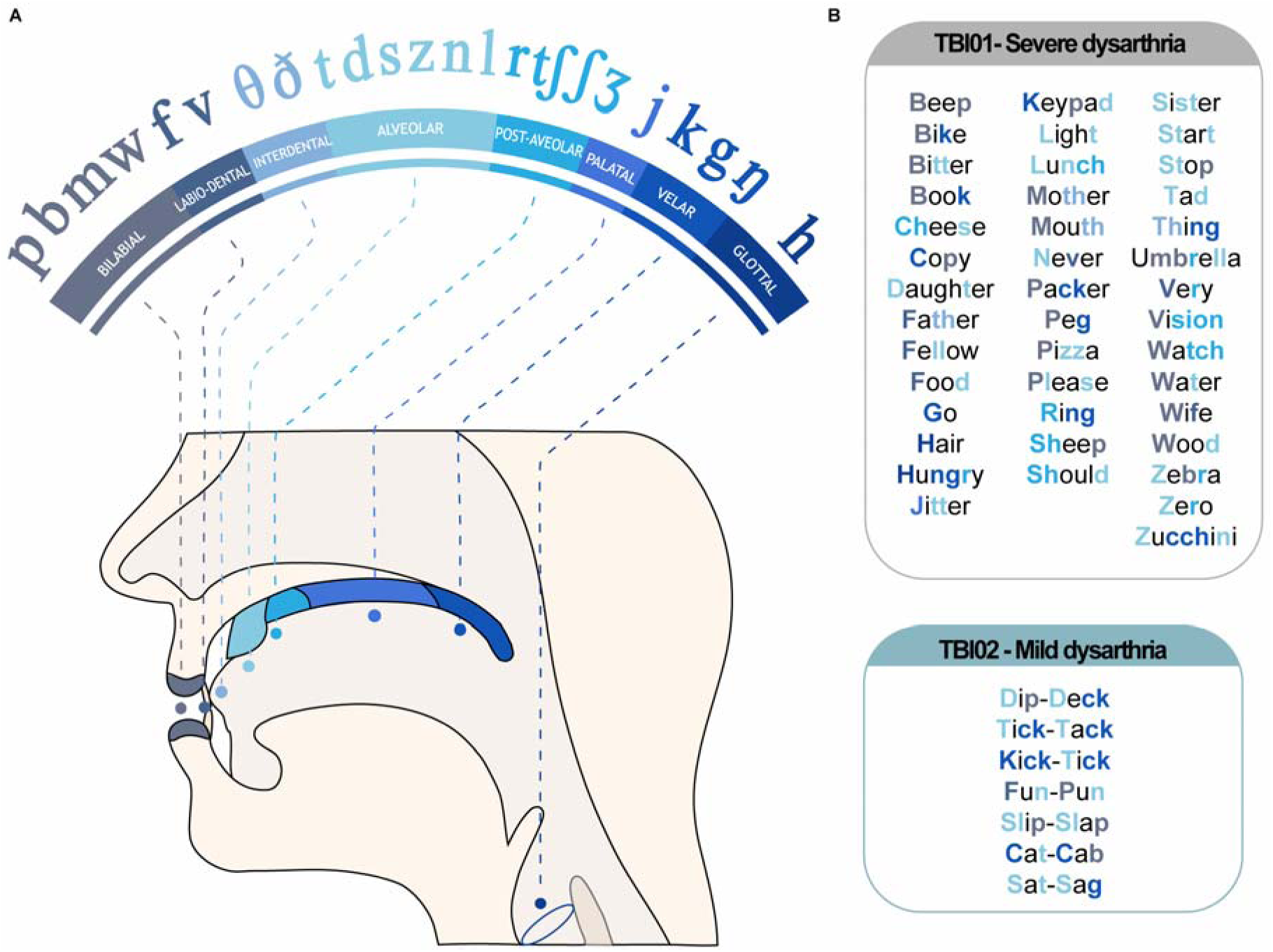
Words tested over the different sessions for TBI01 and TBI02. (a) *Left*: International Phonetic Alphabet schematic^92^ of consonant phonemes distribution. (b) *Right Top*: All words tested for TBI01. *Right Bottom:* All words tested for TBI02. For both tables, consonant phonemes within each word color coded corresponding to the International Phonetic Alphabet schematic.

**Extended Data Figure 6.**
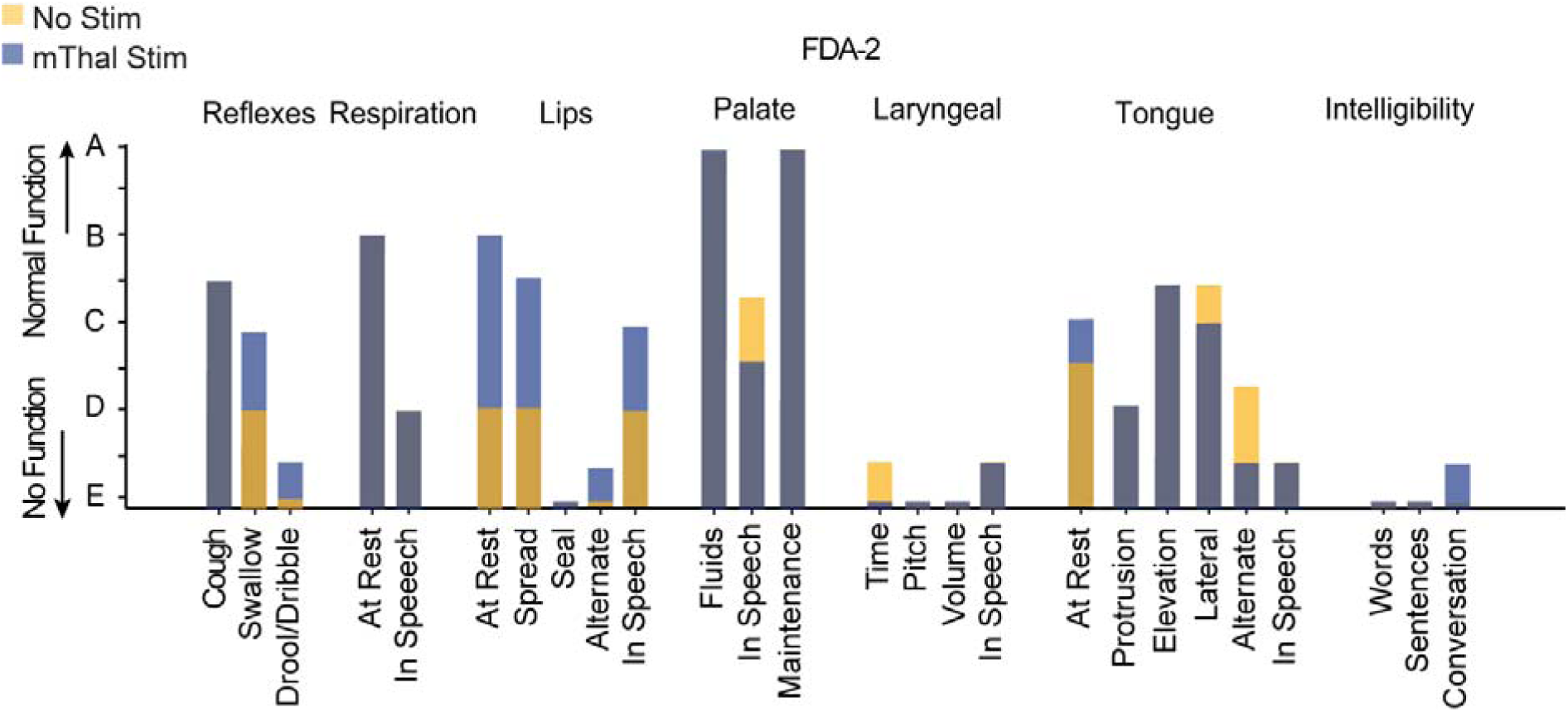
Frenchay Dysarthria Assessment-2 of TBI01. FDA-2 results of TBI01 conducted by blinded SLP for DBS motor thalamus stimulation OFF (yellow) and ON (blue) conditions. For each item, bars are overlapped between the two conditions.

### SUPPLEMENTARY TABLES

**Supplementary Table 1:** Summary of clinical details of the participants enrolled in the study and of the behavioral experiments performed. The latter were selected for each participant based on their deficits and time available for experiments. For additional information about CBT and CST damage see **Extended Data Figure 3**. For additional details about speech deficits for TBI01 and TBI02 see Results and Methods.

| Subject | Lesion damage | Speech and swallowing deficits | Experiments performed |
| --- | --- | --- | --- |
| S01 | – | – | DCS MEPs |
| S02 | – | – | DCS MEPs |
| S03 | – | – | DCS MEPs |
| S04 | – | – | DCS MEPs |
| S05 | – | – | DCS MEPs |
| S06 | – | – | DCS MEPs |
| S07 | – | – | DCS MEPs |
| S08 | – | – | Facial expression tasks |
| S09 | – | – | Facial expression tasks |
| S09 | – | – | Facial expression tasks |
| TB101 | Bilateral CBT and CST damage | Profound dysarthria<br>Moderate dysphagia | Facial expression tasks<br>Videofluoroscopic swallow study<br>Speech tasks |
| TB102 | Unilateral CBT damage | Mild dysarthria<br>No dysphagia | Speech tasks |

**Supplementary Table 2:** Stimulation parameters for M1-DCS and motor thalamus stimulation in participants undergoing DBS implantation (S01-S07). The amplitude of stimulation of the motor thalamus was always at 4mA and the pulse width at 100us.

| Subject | Etiology | DCS intensity | Motor thalamus frequencies tested | Facial Muscles showing clear MEPs with DCS alone |
| --- | --- | --- | --- | --- |
| S01 | ET | 9 mA | 50 Hz, 100 Hz | Mass/Mylo/Oris |
| S02 | PD | 2 mA | 50 Hz, 80 Hz | Mass/Mylo/Oris |
| S03 | ET | 8 mA | 50 Hz, 80 Hz | Mass/Mylo/Oris |
| S04 | ET | 11 mA | 50 Hz, 80 Hz | Mass/Mylo/Oris/Crico |
| S05 | PD | 6 mA | 50 Hz | Mass/Mylo/Oris/Ment/Crico |
| S06 | ET | 11 mA | 50 Hz, 100 Hz | Mass/Mylo/Oris |
| S07 | ET | 13 mA | 80 Hz | Mass/Mylo/Oris/Ment |

**Supplementary Table 3:** Motor thalamus stimulation parameters for subjects S08-S10 and TBI01-TBI02.

| Subject | Stimulation Parameters | Facial Muscles |
| --- | --- | --- |
| **So8** | 1 mA, 100 Hz, 100 us | Mass/Mylo/Oris |
| **So9** | 2.5mA, 100 Hz, 100 us | Mass/Mylo/Oris |
| **S10** | 1mA, 50 Hz, 100 us | Mass/Mylo/Oris |
| **TBl01** | 2-4mA, 55Hz, 90-100 us | – |
| **TBl01 (130 Hz)** | 2.0mA, 130Hz, 100us | – |
| **TBl02** | 1.5-5.5mA, 50 Hz, 100us | – |

**Supplementary Table 4:** Behavioral tests analyzed broken down by sessions for TBI01 and TBI02.

| **Subject** | **Session** | **Tasks** |
| --- | --- | --- |
| **TBl01** | **Session 1** | Single Words |
|  | **Session 2** | Single Words |
|  | **Session 3** | Single Words |
|  | **Session 4** | Facial expression tasks |
|  | **Session 5** | Single Words,<br>Passage Reading,<br>Two-word phrases |
|  | **Session 6** | Facial expression tasks,<br>Single Words,<br>Two-word phrases |
|  | **Session 7** | Single Words,<br>Sentences |
|  | **Session 8** | Single Words,<br>Sentences |
|  | **Session 9** | Facial expression tasks,<br>Single Words |
|  | **Session 10** | Assessment of Intelligibility of<br>Dysarthric Speech (AIDS) |
|  | **Session 11** | Assessment of Intelligibility of<br>Dysarthric Speech (AIDS) |
|  | **Session 12** | Assessment of Intelligibility of<br>Dysarthric Speech (AIDS) |
|  | **Session 13** | Frenchay Dysarthria Assessment-2<br>(FDA-2) |
|  | **Session 14** | Videofluoroscopic swallow study |
| TBlo2 | Session 1 | Tongue Twister,<br>Spontaneous conversation |
|  | Session 2 | Tongue Twister,<br>Spontaneous conversation |

## Notes

### Funding Statement

TThe study was executed through the support of internal funding from the Department of Physical Medicine and Rehabilitation at the University of Pittsburgh to EP and from the Department of Neurological Surgery at the University of Pittsburgh to JGM. Additionally, JGM discloses support for the research described in this study from the Hamot Health foundation and the National Institute of Health (R01NS122927). BZM was supported by R01EY028535 and R01NS089069.

